# A Systematic Review of Literatures: To Identify Non-Cancer Drugs Repurposed for Lung Cancer

**DOI:** 10.1101/2023.12.03.23299334

**Authors:** Saikat Samadder

**Affiliations:** Department of Pharmacology, Yonsei University College of Medicine, 50 Yonsei-ro, Seodaemun-Gu, Seoul, Korea

**Keywords:** Drug repurposing, drug repositioning, lung cancer, clinical trials, randomized controlled trials

## Abstract

**Aim:** The aim of this study is to identify literature suggesting drug repurposing based on various medicines found to possess enhanced efficacy against lung cancer in humans based on pre-clinical and retrospective analysis. Often pre-clinical and retrospective study findings may deviate from the results of randomized prospective clinical studies, this study intends to identify the efficacious new drugs against lung cancer in placebo-controlled studies.

**Methods:** A systematic review of literature was performed in PubMed database on 30^th^ August 2023 without any fixed timeline with terms related to ‘drug repurposing’ and ‘Lung cancer’. Post screening of literature, the names of non-chemo drugs identified were further searched in clinicaltrials.gov additionally NCT# was searched in google scholar, in-case the trial results were not available in respective source to validate the study findings.

**Results:** In PubMed 513 articles were found, 9% articles mentioned about drugs with potential efficacy against lung cancer in humans. In these 45 articles 61 non-chemo drugs were identified from specific 293 clinical studies. Among these 30 studies for 23 drugs were in advanced clinical stage and 9 non-chemo medicines were identified to have positive results in randomized controlled clinical trials, rest of the drugs failed to attend designated clinical efficacy till-date. Anti-alcoholism, anti-bacterial, anti-emetics, cox-2 inhibitors, and HMG-CoA reductase inhibitor are the drugs that could potentially effective against advanced non-small cell lung cancer.

**Conclusion:** Applying the advanced search methodology to identify drugs repurposed or approved for targeted disease. All these 9 non-chemo were screened under randomized trial and currently only one drug disulfiram was found to enhance survival and response, reduced disease progression, in advanced NSCLC originally indicated for anti-alcoholism.

## Introduction

Drug repurposing and repositioning are two similar terms often used to define the reuse of approved candidate drug for a completely new indication [1, 2]. For example, anti-viral (zidovudine) repurposed for cancer, and treatment of HIV infection, at the same time analgesics (aspirin) for relieving pain, and anti-coagulant are well known as repurposed medicines (3). Drugs such as 5-flourouracil, and doxorubicin hydrochloride could be considered as repositioned or redirected drugs prescribed to treat multiple types of blood cancer [2]. Drug manufacturing companies spend millions of dollars to identify new pathways or mechanisms by performing proof-of-concept based studies that could mitigate a diseased condition in patients different from originally indicated disease is better defined as drug reprofiling or rediscovery [4]. Considering, that each anti-cancer drug cannot be prescribed for a particular type of cancer as drugs are always cancer or stage specific, due to drug resistance a drug may not be capable of reducing tumor burden in populations, these facts create never ending scope of developing the most effective, efficient, and efficacious drug. There are 1248 drugs approved by USFDA, there are more than 270 anti-neoplastic agents dedicated for cancer treatment [5, 6]. Meanwhile, lung cancer is the second most commonly diagnosed cancer in the world after breast cancer. As per GLOBOCAN report of 2020, globally 2.2 million new cases are reported per year and more than 1.7 million deaths occurs yearly due to lung cancer [7].

In real-world settings patients are treated with more than 900 types of non-chemo medicines to reverse diseased conditions such as fluid accumulation, anorexia, cachexia, various infections, nausea, vomiting, pain, organ failure, immunosuppression, and hyperactivation of immune system caused by anti-neoplastic agents or cancer. Therefore, often real-world evidence (RWE) is systematically reviewed with retrospective study design to identify possible disease modifying effects by non-cancer drugs mainly increased overall survival (OS), disease-free survival (DFS) in patients suffering from various comorbidities [1, 2]. The actual prescribed indication does not vary from the original approved indication. This allows an analysts or physicians to rediscover the disease mitigating effect in patients pre-diagnosed with more severe diseases such as cancer [8]. Drugs such as metformin, statins, anti-depressants, and aspirin etc. were found to have disease modifying effects in lung cancer patients found in retrospective observational studies [9, 10]. This results of mechanism of action (MOA) or proof of concept studies are performed to provide evidence for retrospective analysis [11]. However, in prospective or randomized placebo-controlled trials the result always varies from the retrospective study or pre-clinical studies [12].

The clinical trials are designed to validate the efficacy and/or safety of a non-chemo drug, often fails to demonstrate therapeutic efficacy when prescribed as a single agent, whereas in presence of chemotherapeutic or anti-neoplastic agents a drug often outperforms in patient group treated with only anti-neoplastic drug. This does not hold sufficient ground for regulatory authorities to approve it as repurposed drug unless validated under placebo-controlled trials [13]. Meanwhile, in clinical trials drug-drug interaction, OS, and drug safety are primary focus of randomized prospective studies applying non-chemo medications for new indication such as non-small cell lung cancer (NSCLC) or any other life-threatening diseases. The findings of a study with review of all approved drugs within a specific time-frame found that there was little or no difference in the evidence supporting supplemental and original indicated approvals, as the number and design of pivotal trials supporting supplemental indicated approvals never lacked well-controlled study designs [14]. USFDA drug policy on drug repurposing the 21^st^ century cures act of 13^th^ December 2016; suggests approval of a drug for cancer, or any other disease based on study design, most rarely RWE is utilized to support the clinical trial with a historical placebo control arm designed by propensity score matched within the study groups. There was huge applause among researchers regarding metformin for treatment of cancer. However, adding metformin to chemotherapy for advanced NSCLC was safe but did not significantly improve clinical outcomes compared to historical phase 3 control group [15]. Recently, the largest phase 3 randomized trial of metformin adjuvant therapy for breast cancer, enrolled 3,649 women with a 5-year follow-up, found no benefit for DFS or OS treated with metformin. Failure to produce clinical efficacy and application of placebo-controlled in trial are not the only reason for disapproval [13]. The failure of a drug in phase trials is also related to two major types of biases produced in retrospective analysis patient selection and time related biases [12].

Based on retrospective analysis with several deficits in study design suggests drug repurposing supported by pre-clinical trials, here it will be further evaluated that the clinical success of non-chemo medicines in randomized double-blinded placebo-controlled trials. The main purpose of this study is to identify the non-chemo drugs suggested to potentiate anti-neoplastic effect in patients with lung cancer. Here it is further validated the clinical study types, phase, its adverse events, response rate, safety and efficacy results to identify the extent of drug’s ability to provide support lung cancer patients at low cost. Including its approval status, and extent of current clinical application are demonstrated here. Lastly, the non-chemo drugs capable of reducing cancer burden reached to clinical trial stage will be identified by pre-clinical or retrospective study type and to understand the clinical success rate contributed by these approach as supporting evidence adopted or encouraged clinical phase trial. Till date there is no study available to suggest most potential non-chemo drug supposed to be repurposed for lung cancer patients based on evidence of clinical trials.

## Methods

### Primary Search strategy

The preferred reporting items for systematic reviews and meta-analyses (PRISMA 2020) statement for reporting was utilized [16]. PubMed database was searched on 30^th^ Aug 2023 with the following terms ((Drug repurposing) OR (drug repositioning)) OR (drug rediscovery) OR (drug redirecting) OR (drug reprofiling) AND (lung cancer)) without any start timeline applied, to identify all the relevant literatures available [2]. Utilizing these terms as search strings can be considered sufficient as these terms accurately identified articles published in the recent past. The articles found in PubMed were excluded or included based on specific criteria, are provided in (**Figure 1**). Primarily, duplicates and non-English, inaccessible articles were excluded. Secondary, exclusion was applied on articles mainly focusing on in-vitro and in-vivo models, in-silico studies, studies on COVID-19, pulmonary hypertension or fibrosis were excluded. Lastly, studies or review articles reporting or suggesting non-chemo drugs for lung cancer based on findings in human clinical trials were included. Chemo drugs already indicated for the treatment of lung cancer were excluded, few originally indicated for infections later used for cancer treatment but not indicated for lung cancer were considered for inclusion. One study on COVID-19 with patients comorbid with lung cancer suggested doxycycline repurposing was included because another pre-clinical study provided strong evidence (**supplementary material**).

**Figure 1:**
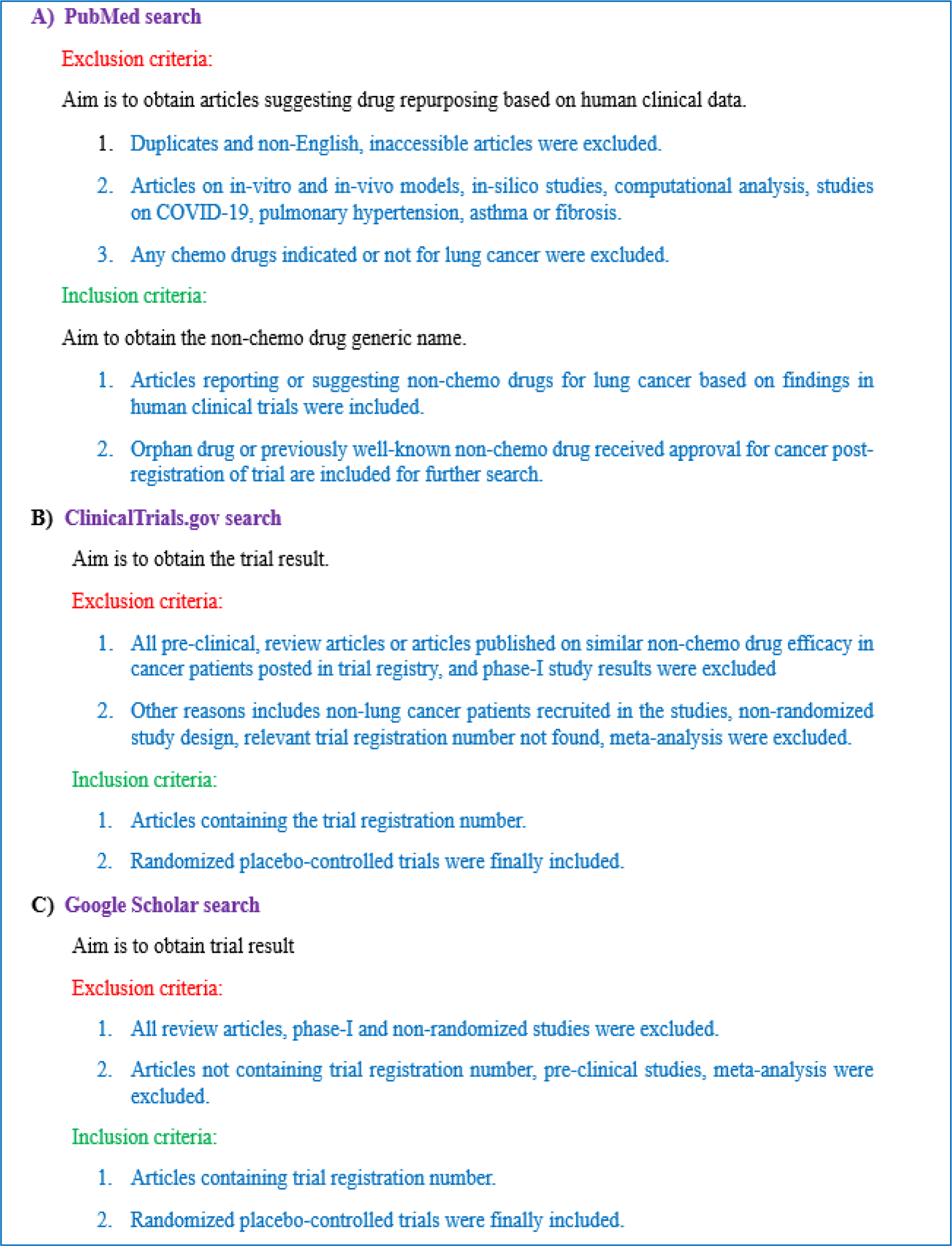
Inclusion and exclusion criteria.

### Additional Search Strategy and Results

The search fetched a total of 513 literatures, 2 articles were found to be duplicates, 2 articles were inaccessible, 1 article was found as non-English. Records screened 508 among these 73 articles were not related to lung cancer, among these 435 articles were found to contain findings not related to lung cancer. These 435 articles were sought for retrieval and 48 articles on SARS-CoV-2 related were excluded, 342 articles were pre-clinical studies, in-silico drug discovery studies or computational analysis, pulmonary hypertension, asthma, and related reviews or studies not referring to clinical trials were excluded. Remaining, 45 articles or reviews reported various chemo and non-chemotherapeutic agents for lung cancer in humans with strong clinical evidence was suggested (**supplementary Table S1**). In these 45 articles reported 84 different non-chemo drugs were under phase trials for repurposing against various types of lung cancer no published results (**supplementary figure S2**).

In second step each “non-chemo” drug name and disease ‘‘lung cancer’’ as combination were used to search clinicaltrials.gov on or before 31^st^ October 2023. There were 23 drugs out of 84 were found with no studies available in ClinicalTrials.gov were excluded (**supplementary table S3**). Total, 61 out of 84 non-chemo drugs were found with or without trial results for repurposing, and remaining 284 trials were identified and included (**supplementary table S4**) for 61 drug candidates were further searched in clinicaltrials.gov for trial identifier and to evaluate the status, and available literatures and the trials specifically designated and designed to validate the therapeutic efficacy in patients with various patients of lung cancer. Often the trials registered in clinicaltrials.gov for non-chemotherapy drugs for lung cancer lacked the study results with published article title (n = 117), irrelevant articles (n=751), and phase-I study result and other reasons (n=35) were excluded. From 47 articles 12 articles and two NCT03861091, NCT00527319 for 14 non-chemo drugs were in phase II/III clinical studies from clinicaltrials.gov were included. The criteria of inclusion and exclusion for articles in clinicaltrials.gov are shown in (**Figure 1**).

Considering insufficient number of articles found, with the trial registration numbers (n=293 NCT#) were further searched in Google Scholar or before 31^st^ October 2023 to identify the findings published as abstract in conferences or research studies, in some cases, systematic review, and meta-analysis (n = 1110) reported for a specific non-chemo drug for lung cancer were excluded, only trial identifier available in published article text were considered and included for further review. A total of 89 clinical trial results were found (n=32) abstracts, (n=12) phase-I studies, and (n=16) articles excluded for other reasons mainly NCT# was not available in the text additional (n=7) articles non-randomized were excluded. Finally, (n=19) research articles contained published study results from randomized blinded or open-label placebo-controlled trials for (n=22) non-chemo were available for inclusion from google scholar, these articles reported only OS, adverse events, safety, and efficacy of 23 non-chemo medicines were finally included from clinicaltrials.gov or google scholar search. Abstracts published for phase-II/III trials were excluded due to risk of reporting bias. There were 7 articles published articles on 6 non-chemo drugs phase-II trials performed in non-randomized open-label trials were excluded from final assessment but trial results are shown in **supplementary table 5**. Total, 28 articles plus 2 trial results obtained directly from (NCT03861091, NCT00527319) clinicaltrials.gov reported about non-chemo drug’s (n=23) clinical success or failure in published or unpublished form (**table 1**). The criteria of inclusion and exclusion for articles found in google scholar are shown in (**Figure 1**).

**Table 1:** Phase II/III trial with randomized double blinded placebo-controlled trials, or randomized double-blinded study findings of 30 studies on advanced non-small cell lung cancer patients treated with chemo, immunotherapy or radiotherapy with 23 non-chemo drugs from pivotal studies are presented. SBRT; Stereotactic Body Radiation Therapy, OS; Overall Survival, PFS; Progression Free Survival, ORR; Overall response rate, LC; Lung Cancer,

| **Drugs (Clinical Phase)** | **Trial identifier** | **Drugs** | **Clinical findings** | **Ref** |
| --- | --- | --- | --- | --- |
| Disulfiram (phase-II) | NCT00312819 | Cisplatin, vinorelbine plus placebo or disulfiram | Significantly higher PFS and OS in disulfiram group compared to chemo plus placebo group. Quality of life and response rate was not statistically significant. | 59 |
| Metformin (Phase-II) | NCT01864681 | Gefitinib with placebo or metformin | No significant difference in ORR and PFS between the groups. Addition of metformin showed 22 months survival and higher incidence of diarrhea, compared to 27 months overall survival in gefitinib group with reduced diarrhea. | 48 |
| Minocycline (phase-II) | NCT00473083 | Erlotinib with delayed or early minocycline treatment | Early minocycline exposure reduced severe incidence of rash compared to delayed exposure patients with no major difference in survival. | 35 |
|  | NCT01636934 | Post-chemo therapy with or no minocycline | Reduced grade 3 adverse events pain, fatigue, and shortness of breath in minocycline group compared to placebo. | 36 |
| Pravastatin (phase-III) | NCT00433498 | Carboplatin, cisplatin, etoposide and/or pravastatin | No significant difference was found for pravastatin compared to placebo in SCLC subjects for OS, and PFS. | 19 |
| Aprepitant (phase-III) | NCT02161991 | Palonosetron & Dexamethasone with or Without Aprepitant. | Post chemo aprepitant exposure significantly reduced vomiting but not nausea compared to unexposed patients. | 60 |
| Aspirin (phase-III) | NCT02123849 | Low dose intermittent Vs continuous aspirin exposure. | No significant difference in adverse events observed between groups. Continuous exposure showed smoking associated gene expression pattern. | 41 |
|  | NCT02169271 | Low dose aspirin or placebo | There was no significant negative change in the size of the tumors in patients of the treatment group compared to placebo. | 42 |
| Zoledronic acid (phase-III) | NCT00172042 | Vit. D and calcium supplementation with or without zoledronic acid | PFS, OS, and bone metastasis along with renal adverse event were not significantly different among groups. | 56 |
|  | NCT00264420 | Zoledronic acid infusion or placebo in lung cancer patients. | Significantly reduced and delayed skeletal related events in patients treated with zoledronic acid, bone pain was observed in both groups. | 54 |
| Denosumab (phase-III) | NCT01951586<br>NCT02129699 | Chemotherapy-denosumab and chemotherapy-alone | Denosumab group did not show significant increase in OS, PFS. In subgroup analysis denosumab performed better than chemo alone in squamous cell carcinoma patients. | 53 |
| Thalidomide (phase-III) | NCT00004859 | Carboplatin, paclitaxel, radiation and/or thalidomide. | Inclusion of thalidomide severely affected OS and grade 3 adverse events. | 30 |
|  | NCT00061919 | Etoposide, carboplatin placebo or thalidomide | No difference in OS, and PFS. All severe and non-severe adverse events were more frequent in thalidomide group. | 29 |
| Rofecoxib (phase-III) | NCT00385606 | Rofecoxib plus gemcitabine, and cisplatin | Rofecoxib did not prolong OS, but improved PFS, ORR, reduced adverse events mainly pain, and quality of life. | 43 |
| Celecoxib (phase-III) | NCT01041781 | Carboplatin plus gemcitabine for squamous and carboplatin plus | No significant difference in OS, PFS treated Vs placebo among groups. Significant difference in OS and PFS was observed post separation of patients based on high | 44 |
|  |  | pemetrexed for non-squamous NSCLC with celecoxib or placebo | prostaglandin E2 metabolite from baseline. |  |
|  | NCT00300729 | Palliative chemotherapy with celecoxib or placebo | No significant difference in OS, quality of life and pain was observed in celecoxib group compared to placebo. Females were more benefited from celecoxib than males in terms of OS. Leukocytopenia was frequent in celecoxib patients. | 45 |
| Apricoxib (phase-II) | NCT00652340 | Erlotinib with Apricoxib or alone with placebo | Time to disease progression was found increased in apricoxib patients <65 years old. | 46 |
| Carboxyaminoimidazole (Phase-III) | NCT00003869 | Post chemotherapy carboxyaminoimidazole or placebo | No significant difference in OS or disease progression observed between groups. Significantly lesser adverse events in placebo. | 39 |
| Itraconazole (phase-III) | NCT03664115 | Platinum-based chemo or with itraconazole | The ORR and PFS are significantly higher in itraconazole group compared to placebo but no difference in overall survival. | 31 |
| Risedronate (Phase II) | NCT03861091 | Post SBRT risedronate dosing | Reduced bone loss in lung cancer patients compare to placebo. No serious adverse event occurred in patients of treatment group. | 54 |
| Retinoic acid (Phase II) | NCT00002586 | 13 cis retinoic acid with or without tocopherol | No significant change observed in positive histological changes compared to placebo patients. | 40 |
| Etodolac plus propranolol (VT-122) (phase II) | NCT00527319 | Post-surgery VT-122 or placebo | Post 1 month of therapy no major change in body mass or griping strength, 25% severe adverse events seen in treatment groups. | 18 |
| Simvastatin (Phase-II) | NCT00452244 | Gefitinib alone or with simvastatin | Significant statistical difference seen in ORR, PFS. The OS was 13.5 months and 12 months in gefitinib plus simvastatin and gefitinib placebo groups, respectively. | 20 |
|  | NCT01156545 | Afatinib alone or with simvastatin | The ORR, PFS and OS was not significantly higher in Afatinib plus simvastatin group. Skin rash was mostly seen in Afatinib group. | 21 |
| Pioglitazone and clarithromycin (Phase-II) | NCT02852083 | Nivolumab plus treosulfan, pioglitazone, clarithromycin or nivolumab and placebo. | No statistical significance observed in PFS, ORR, OS, grade 3-5 adverse events was higher in nivolumab group. | 28 |
| Pioglitazone (Phase-II) | NCT00780234 | Pioglitazone alone or placebo in former or current smokers with high-risk LC subjects. | Pioglitazone did not improve dysplasia in patients. | 49 |
| Nitroglycerine (Phase II) | NCT01171170 | Carboplatin, bevacizumab, paclitaxel with or without nitroglycerine. | Nitroglycerine did not improve OS and PFS compared to placebo. Headache was most prominently observed in treated patients. | 22 |
|  | NCT04338867 | Nitroglycerine plus whole brain radiotherapy (WBRT) or WBRT alone. | Nitroglycerine arm had enhanced ORR, PFS but not OS compared to placebo arm. | 23 |
| Doxycycline (Phase-II) | NCT00531934 | Erlotinib alone or doxycycline combined | Reduced severity of folliculitis in doxycycline arm compared to control group patients. | 38 |
|  |  | Post-dacomitinib | Significantly reduced dermatological events and increased |  |
|  | NCT01465802 | treatment, doxycycline, placebo. | quality of life in doxycycline treatment group compared to placebo. | 37 |

Articles identified as randomized trials were primarily included but based on availability of randomized placebo-controlled trial result irrespective of negative or positive results, the study was included and any other publications related to same drug reported based on non-randomized, open-label, phase-I studies were not included in final analysis or in **table 1**. In-case two studies on same drug with different treatment plan and drug was available then both the studies were presented, reports removed based on strength of design, reporting, and selection biases. Each of the steps to identify literatures was carried twice to ensure reliability of data assessed independently by a first author further confirmed by corresponding author. The non-chemo drugs list, indication, approval year/status, reference from primary search approach that led to phase trial, present status of candidate drug and availability or unavailability of publication for respective drug is presented in tables (**supplementary table 1 & 2**). The numbers of articles, non-chemo drugs, and clinical trials included and excluded are provided within PRISMA 2020 flow diagram for new systematic reviews which included searches of databases, registers and other sources (**PRISMA flow chart 1**).

Three drugs eprenetapopt, romidepsin, and arsenic trioxide were considered for search in clinicaltrials.gov despite being chemotherapeutic agents because the trials started prior to approval for respective oncogenic indications but were excluded from final assessment. All drugs list suggested in various studies obtained from 423 articles is presented in a table with 57 non-chemo to confirm that clinical trials are not available till-date (**supplementary table 6**). Enalapril maleate, and mirtazapine were found from drugs not reported in pre-clinical studies and not referring human subjects were excluded in final assessment but shown in because ongoing clinical trials available (**supplementary table 4**) and for future tracking of clinical results. Hydroxychloroquine, arsenic trioxide, and nelfinavir were studied in non-randomized trials were excluded from final assessment. Lastly, a bias assessment table is provided and performed similar to constable et al. 2022 [17] with assessment scores is provided in (**supplementary table 7**). There are 6 articles duplicates in clinicaltrials.gov and google scholar highlighted in (**supplementary table 8**). Complete list of 89 articles from google scholar and 47 articles from clinicaltrials.gov are listed in (**supplementary table 9 & 10**). There was no scope of performing meta-analysis due to low number of phase II/III trials available for each drugs. This review was not registered due to the study question as its aim was to identify clinical safety, efficacy and adverse events of non-chemo drugs applied concomitantly or post chemo-chemo therapy in lung cancer patients. Articles in clinical benefit section could be considered as final analyses and not the conclusion.

## Results

### Cardioprotective Compounds

Anti-hyperlipidemia medications such as simvastatin, pravastatin, anti-hypertensive drugs propranolol, and nitroglycerine prescribed for angina were found effective in pre-clinical studies in in-vitro and in-vivo cancer models. These drugs were studied in advanced lung cancer patients. Propranolol plus etodolac (VT-122) combined treatment was considered for cachexia post radiotherapy. Combinatorial usage did not induce sufficient therapeutic benefit in patients with lung cancer in terms of grasping and weight gain [18]. Pravastatin was widely studied in-vitro and was found to block 3-hydroxy-3-methyl-glutaryl-coenzyme A (HMG-CoA) reductase thereby regulating cholesterol biosynthesis in tumor cells. In a phase-III study design with performance status 0-3 of patients randomly assigned, pravastatin did not extend OS or progression free survival (PFS) compared to placebo [19]. Simvastatin combined with gefitinib or afatinib was studied in two separate well-controlled trials suggested that gefitinib and simvastatin together were capable to prolong survival and PFS in advanced NSCLC patients, but similar efficacy was not observed in treatment group of afatinib and simvastatin compared to control group [20, 21]. Anti-angina medication nitroglycerine was trailed with other chemotherapeutic agents such as carboplatin, bevacizumab, paclitaxel with/without nitroglycerine did not achieve sufficient safety or efficacy compared to placebo [22]. In a study randomly including stage-IV NSCLC brain metastasis patients treated with whole brain radiation therapy and nitroglycerine or WBRT alone found increased PFS and ORR in nitroglycerine group compared to placebo. There was no difference in OS in nitroglycerine treatment group compared to placebo [23]. In meta-analysis of randomized controlled studies and observational studies separately conducted on of lung cancer patients treated with statins found increased OS [24, 25].

### Anti-microbial agents

There are numerous anti-microbial and anti-parasitic agents available for the treatment of infections. Among those drugs such as thalidomide, itraconazole, minocycline, doxycycline, clarithromycin, arsenic trioxide, was frequently studied in pre-clinical settings. Thalidomide was indicated for treatment of leprosy (erythema nodosum leprosum), later used for the treatment of graft versus host diseases, in 1998 it was approved for multiple myeloma [26]. Arsenic trioxide was previously indicated for the treatment of syphilis, in 1994 it was approved for the treatment of acute promyelocytic leukemia [27]. The non-randomized study on arsenic trioxide is not presented here. Clarithromycin was not studied as single agent in clinical trials and was studied in presence of pioglitazone and treosulfan [28]. Clinically thalidomide, minocycline, doxycycline was evaluated under randomized, double-blind, placebo-controlled study design.

In RCT’s with thalidomide 724 SCLC patients with thalidomide or placebo were further evaluated and found no significant difference in-terms of OS, and quality of life among groups [29]. In second study including advanced NSCLC patients were treated concomitantly with paclitaxel, carboplatin and 60 Gy radiation and thalidomide or placebo. There was no difference in OS, PFS, and ORR among two groups. Incidence of grade 3 or 4 adverse events were higher in thalidomide group compared to placebo [30]. Itraconazole was evaluated under randomized open-label placebo-controlled trial including chemo naïve advanced NSCLC patients treated with platinum-based chemo plus itraconazole regularly for 21 days or placebo. There was significant ORR, and PFS among groups but no difference in 1-year OS [31]. Anti-bacterial medications such as minocycline and doxycycline were found to be inhibit metalloproteinase and vascular endothelial growth factor (VEGF) reducing angiogenesis and degradation of extracellular matrix (ECM) [32, 33, 34]. In a randomized clinical study advanced NSCLC patients treated with erlotinib with delayed or early minocycline or placebo found early minocycline exposure reduced severe incidence of rash compared to delayed exposure patients with no significant difference in OS [35]. In another study post-chemo therapy with minocycline or alone successfully reduced grade 3 adverse events in minocycline group compared to placebo [36]. Post-dacomitinib treatment of advanced NSCLC patients randomly received doxycycline or placebo and found to significantly reduce the dermatological events and increased quality of life in doxycycline treatment group compared to placebo [37]. In an open-label RCT erlotinib alone or doxycycline combined reduced severity of folliculitis in doxycycline arm compared to control group patients [38]. In an RCT post chemotherapy carboxyaminoimidazole or placebo was applied on patients to evaluate the efficacy and safety found that no significant difference in OS or disease progression observed between groups, significantly lesser adverse events observed in placebo arm [39].

### Immunosuppressive drugs

There are 6 known anti-inflammatory drugs were tested in lung cancer patients, such as aspirin, celecoxib, apricoxib, Rofecoxib, etodolac and 13-cis-retinoic acid. The drug 13-cis-retinoic acid indicated for acne and all-trans retinoic acid is approved for acute promyelocytic leukemia. Randomly selected lung cancer patients treated with 13 cis retinoic acid with or without tocopherol found no significant change observed in positive histological changes compared to placebo patients [40]. In a randomized double-blinded study aspirin was prescribed to current heavy smokers previously diagnosed with lung cancer, grouped as per dosing continuously or intermittently (one week aspirin and one week placebo). Low dose (81mg) aspirin did not prevent carcinogenesis and incidence of adverse events were similar in both arms [41]. In another study on patients with solid nodules of lungs randomized to receive aspirin or placebo found that there was no significant negative change in the size of the tumors in patients of the treatment group compare to placebo [42]. In 4 different randomized placebo-controlled trials focusing on safety and efficacy of rofecoxib, celecoxib, and apricoxib prescribed along with chemo or immune-therapy regimens found that none of the three anti-inflammatory agents improved OS, rofecoxib and apricoxib improved PFS but celecoxib did not improve PFS in patients compared to placebo [43, 44, 45, 46]. Etodolac and propranolol was combined as (VT-122) was prescribed for cachexia in patients undergoing surgery for lung cancer. After 1 month of therapy with VT-122 no major change in body mass or griping strength, 25% severe adverse events seen in treatment groups [47].

### Anti-diabetic drugs

Anti-diabetic drugs such as metformin was extensively studied in various cancer patients including in advanced NSCLC patients. There are numerous retrospective and prospective studies suggesting clinical benefits such as prolong OS and PFS in lung cancer patients in absence of diabetes. There are very few randomized double-blind placebo-controlled trial with primary focus on NSCLC patients treated with chemotherapy plus metformin or placebo. One such study performed and could not find beneficial effect in patients treated with metformin and gefitinib in terms of OS/PFS or diarrhea compared to placebo [48]. Pioglitazone another well-known anti-diabetic drug prescribed for type-2 diabetes mellitus was studied for clinical efficacy in high-risk lung cancer patients in randomized double-blind placebo-controlled settings with current or former smokers. The results suggested no improvement of bronchial dysplasia in current smoker with minute benefits in former smokers [49]. In another study NSCLC patients failed treatment with platinum-based chemo regimen were included in the study and received treosulfan, pioglitazone, and clarithromycin daily or nivolumab alone. OS of 9.4 months was seen in pioglitazone plus clarithromycin group compared to 6.9 months in nivolumab group, 75% patients in non-nivolumab group required rescue therapy with immunotherapy [28].

### Anti-resorptive drugs

Drugs prescribed for osteoporosis patients such as denosumab, bisphosphonates zoledronic acid, and risedronate [50]. Regular intake of anti-osteoporotic medications post-chemotherapeutic intervention reduced depletion of calcium, commonly seen in patients with metastasis to bone. In a study both denosumab and zoledronic acid induced osteonecrosis of the jaw in patients with 40% lung cancer within the study, hypocalcemia was frequently observed in denosumab arm, while acute kidney related adverse events were seen in zoledronic acid patients. In this study bone related events were more frequent in denosumab group than zoledronic acid treated patients [51]. Similar result was obtained in a study analyzed two randomized trials simultaneously with 740 NSCLC patients treated with chemo plus denosumab and chemo alone. This study reported no significant improvement of OS/PFS in patients receiving denosumab compared to control [52]. On the other hand, study including 773 NSCLC patients treated with zoledronic acid 4 mg regular dosing of zoledronic acid for 21 months or placebo resulted in significant reduction of skeletal adverse events [53]. Although, there is no comparative study available on zoledronic acid Vs risedronate in clinical settings. Interestingly, risedronate potentially decreased bone loss in lung cancer patients compared to placebo group and no serious adverse events occurred in patients of treatment group [54]. Reduced skeletal adverse events could be inferred as reduced disease progression resulting in increased OS but this depends on the frequency, age, gender of patients receiving chemotherapy and/or radiotherapy for advanced stage NSCLC. However, zoledronic acid could not increase OS/PFS, progression of metastasis and impaired renal function in patients treated with zoledronic acid [55].

### Anti-alcoholic agent

There are multiple anti-addiction drugs are available in the market. Among all disulfiram was intensively studied in-vitro and in-vivo cancer models. Disulfiram a metallodrug primary known for inhibiting cellular aldehyde dehydrogenases, it forms copper diethyldithiocarbamate complex (Cu(DDC)_2_ complex) that induces redox reaction by forming Cu(DDC)_2_ resulting in ROS generation causing tumor cell death [56]. Disulfiram is one of the few drugs capable of inducing apoptosis in cancer stem cells by suppressing nuclear factor kappa-light-chain-enhancer of activated B cells (NFκB) pathway [57]. Selective and effective killing of cancer stem cells was linked to increased OS and PFS in advanced NSCLC patients. In multicenter, randomized, double-blinded placebo-controlled study treated advanced NSCLC patients with cisplatin, vinorelbine and disulfiram or no disulfiram, found 10 months OS, and 5.9 months PFS compared to untreated patients survived 7.1 and 4.9 months of PFS. Disulfiram being a regularly prescribed medication for alcoholism has very little-known adverse events caused clinically in patients, further clinical studies demonstrating safety in terms of serious adverse events should be the focus in future [58].

### Neuromodulatory drugs

Desipramine, and valproic acid are currently completed phase-II trials, more studies are required because present studies are open-label single arm studies. Neurokin-1 inhibitor aprepitant completed phase-III trial and was found to achieve desired activity in patients. Aprepitant was prescribed alone or with palonosetron and dexamethasone with or without post-chemo, addition of aprepitant to patient group significantly reduced vomiting but not nausea compared to unexposed patients. Based on these and other advanced study results aprepitant is currently as an approved anti-emetic for chemotherapy-induced nausea and vomiting for cancer patients [59].

### Limitations

The reliability of clinical studies are partially affected due to open-label status that determines the strength of a trial in-terms of biases. Trials those are randomized, double-blind, placebo-controlled design are considered highest standard for identifying drug safety and efficacy. Phase II/III trials recruiting only advanced stage patients accurately generates the clinical end-point results. Evaluation based on compliance in reporting with statistical analyses are efficient in reducing reporting biases. Based on these important facts the drugs found to have potential efficacy and safety such as simvastatin, itraconazole, minocycline, doxycycline, and disulfiram were found beneficial for patients, at the same time these trials contained least overall biases. Minocycline and doxycycline studies were both open and blinded therefore has moderate biases and results are reliable, simvastatin, and itraconazole studies were open label this slightly increases the risk of biases. 13 Out of 30 (50%) studies were performed in randomized double-blind placebo-controlled clinical set-up suggested for non-chemo drug repurposing in advanced NSCLC. The study on disulfiram was a phase-IIb study with very low number of patients, further study is necessary recruiting a larger number of patients. Rofecoxib, apricoxib, celecoxib, simvastatin, itraconazole, and disulfiram including one study related of doxycycline treated patients concomitantly with chemo. Post-chemotherapy drug exposure was found in one study of doxycycline, and two studies of minocycline. The treatment plan were aligned to effectively identify clinical end-points, further reduced reporting bias. Among all 30 randomized trials, the 15 phase-II/III studies with clinical endpoint success are discussed in clinical benefit section as all these recruited a specific type of lung cancer patients, further reducing design, and selection biases. All the trials included in final assessment of this study are related to rofecoxib, apricoxib, celecoxib, simvastatin, itraconazole, minocycline, doxycycline, and disulfiram consisted of low-moderate biases in-terms of study design, patients selection, and reporting. Remaining 15 studies were open label, studies on aspirin lacked placebo arm, two studies on risedronate and VT-122 were unpublished and may contribute to reporting biases. Long term prescribed drugs such as simvastatin or metformin, the accumulated trials consisted of patients non-comorbid with hyperglycemia or hyperlipidemia this reduced selectivity biases. The articles obtained from each searches were eventually excluded are shown in supplementary table thereby reducing biases of this study. COCHRANE database was not searched thereby limiting the studies registered in other geographical locations.

### Clinical Benefits

The primary aim of each of the RCTs was to identify the non-chemo drug’s ability to measure the potentiality of enhancing OS, PFS, and inducing overall response rate (ORR) in advanced NSCLC patients and the trials with high grade study design can effectively suggest drug repurposing are presented in this study. Non-chemo medications such as pravastatin, propranolol, aspirin, etodolac, 13-cis retinoic acid, metformin, pioglitazone, thalidomide, clarithromycin, carboxyaminoimidazole, denosumab, could not achieve desired clinical end-points in the registered trials [18, 19, 28, 29, 30, 39, 40, 41, 42, 48, 49, 53, 54]. Etodolac may not be beneficial for lung cancer but was found effective against colorectal and breast cancer [60, 61]. Similarly, metformin may have more beneficial effect in other cancer than in lung cancer patients. Rofecoxib was found associated with cardiac event in cancer patients compared to placebo patients [62]. Nitroglycerine increased the risk of post-exposure headache in NSCLC patients [22]. Non-chemo such as itraconazole, apricoxib, and rofecoxib could potentially improve ORR and reduce disease progression in patients with NSCLC [31, 45, 46]. Disulfiram along with cisplatin, and vinorelbine was effective in extending survival, response rate, and reduced disease progression in advanced NSCLC patients at the same time simvastatin, and gefitinib in combination would provide similar benefits in advanced stage NSCLC patients [20, 58]. Based on clinical efficacy results of disulfiram in large group of NSCLC patients, it could be repurposed. So far, the safety is concerned simvastatin, minocycline, doxycycline, itraconazole, aprepitant, zoledronic acid, risedronate and rofecoxib contributed to least incidence of adverse events in advanced NSCLC patients [20, 31, 36, 43, 37, 38, 54, 56, 60]. Based on safety and efficacy aprepitant is already indicated for chemo-therapy induced nausea and vomiting in 2003 but it has no effect on OS/PFS [59]. However, reduced nausea increases quality of life in patients undergone heavy or moderate chemo-radio therapeutic regimens. Only study to demonstrate the effect of non-chemo drug (aspirin) in reduction of tumor size failed to find sufficient reduction in tumor size compared to placebo [42]. In a recent review article, found 14 clinical trials designed to screen lung cancer prevention [63]. Compared to our study in a recent systematic review found 11 non-chemo drugs suggested for various cancer patients [64]. The approach utilized here to effectively find RCT is beneficial for obtaining maximum pivotal studies.

### Conclusion

There are more than 150 non-chemo drugs suggested for various cancer treatment [11, 65]. Finding the cheapest drug that could potentially prolong survival in advanced stage patients is of great importance. Currently, none of the non-anti-neoplastic agents are capable of producing tumor killing activity in absence of anti-neoplastic drugs, all the clinical studies presented here were coherent to this well-known treatment plan. In pre-clinical studies single agents produce significant cell deaths of cancerous cell lines, provides sufficient preliminary evidence for further proceeding to in-vivo studies [66]. In retrospective analysis matching concomitant treatment with several other non-chemo drug’s effect is not usually reported by performing propensity score matching resulting in biased reporting. Excluding the fact that in patients with a specific symptom received a particular non-chemo increases a selection biases of retrospective studies [12]. These limits the reliability of evidence provided in retrospective studies for drug repurposing, by using machine learning it could be avoided, although statistical significance similar to placebo-controlled phase-III trials may not be achieved to confirm this further study is required [67]. Nelfinavir is a potential drug candidate capable of producing distinct and advanced tumor destruction with minimal adverse events, it increased OS, PFS, and ORR in patients further well-controlled studies are required in larger population to demonstrate efficacy and safety for the drug to be repurposed [68]. All the studies were performed on advanced stage NSCLC patients, other non-chemo drugs studied in extensive SCLC or malignant methelioma were not effective for repurposing. All the non-chemo successfully reducing adverse events, OS/PFS, were identified pre-clinically, supported by retrospective analysis of RWE, leading to randomized prospective studies to further screen the clinical efficacy, later demonstrated in meta-analysis with efficacy in patient population sub-groups [69]. This flow was followed for all of drugs screened for repurposing, drugs identified from retrospective analysis (aspirin/metformin) may perform well, but distinct clinical efficacy reduces in phase trails was absent. The phase-II/III trials evaluating non-chemo drugs safety and efficacy profile could be the only method for screening therapeutic benefits in patients by conducting advanced clinical trials at the same time historical placebo could be effectively used beside prospectively treatment arm [70]. Simvastatin and disulfiram was found to increase overall survival but phase-III studies are required, recruiting a greater number of patients to obtain distinct clinical benefits. Doxycycline along with bleomycin is prescribed for the management of malignant pleural effusions often reported in patients with various types of lung cancer [71]. Aprepitant significantly reduced several adverse events arising due to chemo/radio therapy such as nausea, pruritus and reduced cough in chemo naïve advanced lung cancer patients [59, 72, 73].

## Supporting information

Manuscript

PRISMA checklist

ICME disclosure

Inclusion and Exclusion criteria

Prisma 2020 flow chart

## Data Availability

All data produced in the present work are contained in the manuscript

## Acknowledgement

The author would like to thank Professor Kyungsoo Park (Yonsei University) for the research questions.

---

**Flow chart 1:**
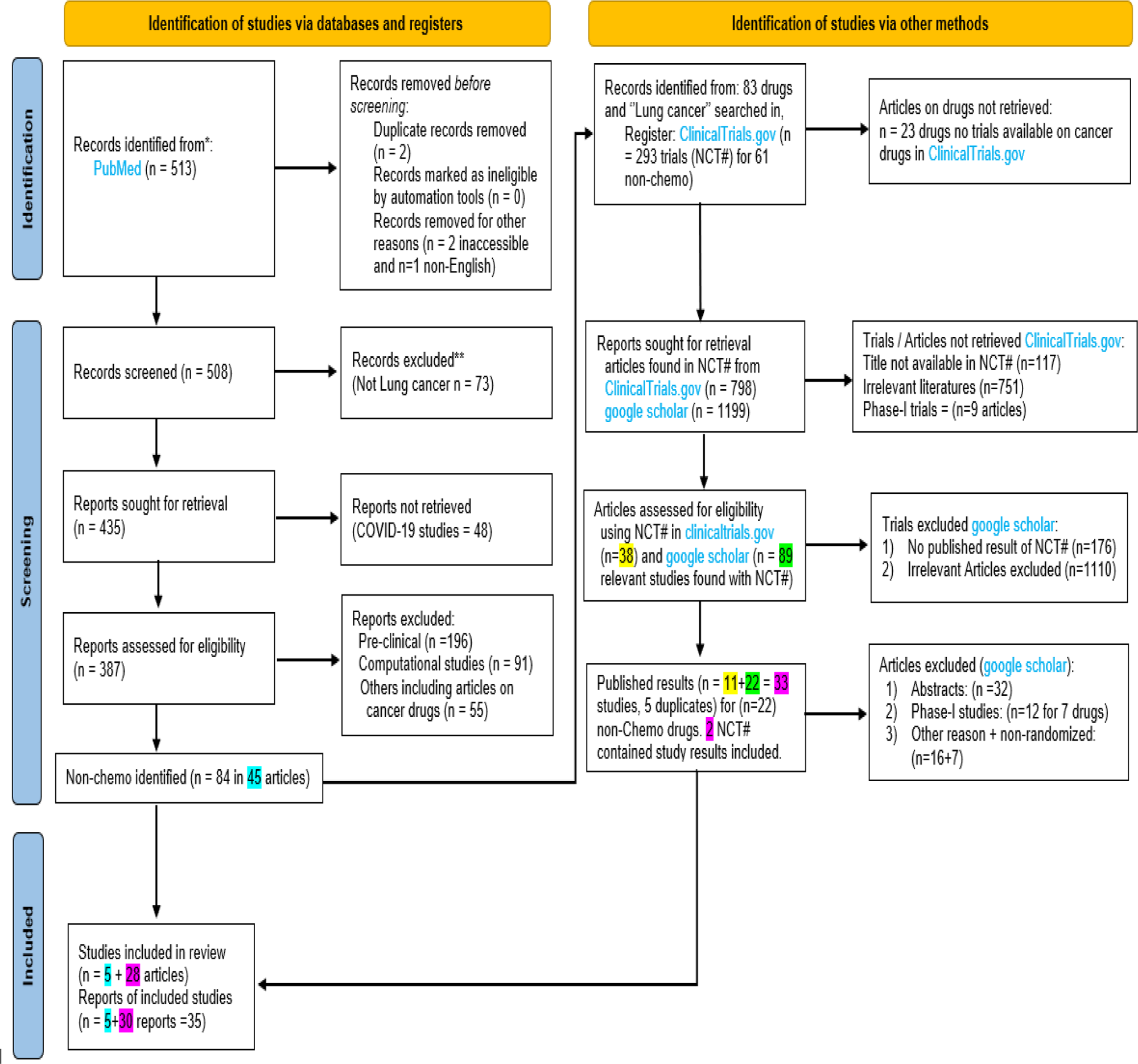
PRISMA 2020 flow diagram for new systematic reviews which included searches of databases, registers and other sources.

