## Supplementary material for "A Systematic Review of Literatures: To Identify Non-Cancer Drugs Repurposed for Lung Cancer": Manuscript

**Contents: Pages**

1. **Supplementary table S1 ----------------------------------------------------- 2-3**

**List of 45 selected articles from PubMed as reference ---------------- 4-8**

2. **Supplementary table S2 ----------------------------------------------------- 9-10**
3. **Supplementary table S3 ----------------------------------------------------- 11**
4. **Supplementary table S4 ----------------------------------------------------- 12-17**
5. **Supplementary table S5------------------------------------------------------ 18**

**List of phase-II non-randomized clinical trials published (reference) - 19**

6. **Supplementary table S6 ----------------------------------------------------- 20-21**
7. **Supplementary table S7 ----------------------------------------------------- 22**
8. **Supplementary Table S8 Combined list of articles obtained from**

**clinicaltrials.gov and Google scholar ------------------------------------- 23-29**

9. **Supplementary Table S9 List of articles from Google scholar ------ 30-35**
10. **Supplementary Table S10 List of articles from clinicaltrials.gov -- 36-38**

| **Drugs** | **Indication** | **Approval year** | **Approval status for Lung cancer** | **Reference from systematic review** | **Initial**  **Approach to reach clinical studies** | **Current phase in Clinical trial** | **Published Results available** |
| --- | --- | --- | --- | --- | --- | --- | --- |
| Disulfiram | Anti-alcoholic | 1949 | Under phase trial | 1, 39, 43 | Pre-clinical | Phase-III completed | Yes |
| Thalidomide | Erythema nodosum leprosum | 1965 | Under phase trial | 4, 6 | Pre-clinical | Phase-III completed | Yes |
| Aspirin | Non-steroidal anti-inflammatory | 1985 | Under phase trial | 4, 6, 7, 8 | Pre-clinical  Retrospective | Phase-III completed | Yes |
| Metformin | Type 2 diabetes | 1994 | Under phase trial | 4, 5, 9, 10, 11, 12, 13, 14, 15, 16, 17, 18, 19, 27, 43 | Pre-clinical Retrospective | Phase-III completed | Yes |
| Itraconazole | Anti-fungal | 1992 | Under phase trial | 1, 5, 6, 9, 10, 12, 20, 21, 43 | Pre-clinical | Phase-III completed | Yes |
| Denosumab | Osteoporosis | 2010 | Under phase trial | 36, 37 | Pre-clinical | Phase-III completed | Yes |
| Minocyclin | Anti-bacterial | 2021 | Under phase trial | 5, 9 | Pre-clinical | Phase-III completed | Yes |
| Simvastatin | HMG-CoA reductase inhibitors | 1998 | Under phase trial | 5, 10, 12, 43 | Pre-clinical  Retrospective | Phase-III completed | Yes |
| Pravastatin | Hyper cholesterolemia | 2000 | Under phase trial | 4, | Pre-clinical & Prospective | Phase-III completed | Yes |
| Celecoxib | Anti-inflammatory | 1998 | Under phase trial | 1, 10, 12, 43 | Pre-clinical | Phase-III completed | Yes |
| Apricoxib | Anti-inflammatory | 2016 | Under phase trial | 10, 12, 43 | Pre-clinical | Phase-III completed | Yes |
| Rofecoxib | Anti-inflammatory | 1999 | Under phase trial | 12, 43 | Pre-clinical | Phase-III completed | Yes |
| Bisphosphonates /  Zoledronic acid | Osteoporosis | 2002 | Under phase trial | 1, 12, 14, | Pre-clinical | Phase-III completed | Yes |
| Aprepitant | Anti-emetics | 2005 | Approved | 25 | Pre-clinical  Prospective | Phase-III completed | Yes |
| Propranolol*  plus  Etodolac  (VT-122) | Anti-hypertension plus  non-steroidal anti-inflammatory drug | 1967 | Under phase trial | 5, 13 | Pre-clinical | Phase-III completed | Yes |
| 13-cis Retinoic acid | Acne | 1982 | Under phase trial | 1 | Pre-clinical | Phase-III completed | Yes |
| Carboxyamidotriazole* | antiparasitic | 1925 | Under phase trial | 8 | Pre-clinical | Phase-III completed | Yes |
| Doxycycline | Antibiotics | 2005 | Under phase trial | 45 | Pre-clinical | Phase-II completed | Yes |
| Clarithromycin | Anti-bacterial | 2000 | Under phase trial | 12 | Pre-clinical | Phase-II completed | Yes |
| Nitroglycerine | Angina (chest pain) | 2000 | Under phase trial | 9, 12, 43 | Pre-clinical | Phase-II completed | Yes |
| Pioglitazone | Type 2 diabetes | 1999 | Under phase trial | 5, 7, 18, | Pre-clinical | Phase-II completed | Yes |
| Risedronate | osteoporosis | 1988 | Under phase trial | 38 | Pre-clinical | Phase-II completed | Yes |
| Arsenic trioxide | Syphilis  Acute promyelocytic leukemia | 1800’s  2000 | Under phase trial | 1 | Pre-clinical | Phase-II completed | Yes |
| Nelfinavir | Anti-viral | 2003 | Under phase trial | 9, 12, 43 | Pre-clinical | Phase-II completed | Yes |
| Hydroxychloroquine | Anti-malarial | 1955 | Under phase trial | 5, 9, 12, 43 | Pre-clinical | Phase-II completed | Yes |
| Desipramine | Tricyclic antidepressant | 1964 | Under phase trial | 23 | Pre-clinical  Retrospective | Phase-II completed | Yes |
| Valproic acid | Anti-seizure | 2008 | Under phase trial | 13 | Pre-clinical | Phase-II completed | Yes |
| Romidepsin | Cutaneous T-cell lymphoma | Approval 2011 withdrawn May 2022 | Under phase trial | 9 | Pre-clinical | Phase-II completed | Yes |
| Quinacrine | Anti-malarial | 2023 | Under phase trial | 9 | Pre-clinical | Phase-I completed | Yes |
| Anastrozole | Non-steroidal aromatase inhibitor | 2000 | Under phase trial | 4 | Pre-clinical | Phase-I completed | Yes |
| Eprenetapopt | Acute myeloid leukemia | 2020 | Under phase trial | 15 | Pre-clinical | Phase-I completed | Yes |
| Olanzapine* | Bipolar disorder | 2004 | Under phase trial | 8 | Retrospective and  pre-clinical | Abstract  Meta-analysis | Yes  Jian-Guo Zhou |
| Atovaquone | Antimicrobial for Pneumocystis jirovecii pneumonia. | 1999 | Under phase trial | 5, 29 | Pre-clinical | Phase-I completed | Yes |
| Rosuvastatin | Hyperlipidemia | 2003 | Under phase trial | 10, | Pre-clinical Retrospective | Phase-I completed | Yes |
| Sirolimus/ rapamycin | Prophylaxis | 1999 | Under phase trial | 11, 22, 35 | Pre-clinical | Completed phase-I trials | Yes |

**Supplementary Table S1:** List of 36 non-chemo drugs suggested for repurposing against lung cancer patients, sequenced as per weightage of evidence. (*) represents the drugs found in clinicaltrials.gov upon search with terms such as selective serotonin reuptake inhibitors, beta-blockers, calcium channel blockers.

**Reference: The 5/45 articles included in main text is highlighted in yellow.**

| **Drugs** | **Indication** | **Approval year** | **Approval status for Lung cancer** | **Reference from systematic review** | **Initial**  **Approach** | **Current phase in Clinical trial** | **Published Results available** |
| --- | --- | --- | --- | --- | --- | --- | --- |
| Perindopril | Hypertension | 1993 | Under phase trial | 9 | Pre-clinical | No study results published (NCT04980716) | _ |
| Metoprolol | Hypertension | 1978 | Under phase trial | 5 | Pre-clinical | No study results published (NCT01281787) | _ |
| Spironolactone | Hyperaldosteronism and hypertension | 1960 | Under phase trial | 5 | Pre-clinical | No study results published (NCT04980716) | _ |
| Trimetazidine hydrochloride | Anti-angina | Approved in France since 1978 | Under phase trial | 5 | Pre-clinical | No study results published (NCT04980716) | _ |
| Atorvastatin | hyperlipidemia | 1999 | Under phase trial | 4, 5 | Pre-clinical | Terminated due to low accrual | _ |
| Nortriptyline | Tricyclic-anti-depressant | 1964 | Under phase trial | 32 | Pre-clinical | No study results published | _ |
| Naproxen | osteoarthritis | 2006 | Under phase trial | 4 | Pre-clinical | No study results published | _ |
| Tocotrienol | Supplement | 2004 | Under phase trial | 1 | Pre-clinical | No study results published | _ |
| Fluoxetine* | Obsessive-compulsive disorder | 1987 | Under phase trial | 8 | Pre-clinical | No study results published | _ |
| Enalapril maleate | Heart failure and chronic hypertension | 1985 | Under phase trial | 44 | Pre-clinical | No study results published | _ |
| Mirtazapine | Insomnia | 1997 | Under phase trial | 9, 44 | Pre-clinical | No study results published | _ |
| Tilarginine | Cardiogenic shock | 2005 | Under phase trial | 41 | Pre-clinical | No study results published | _ |
| Mebendazole | Anthelmintic | 1974 | Under phase trial | 32, 33 | Pre-clinical | Not sure if Lung cancer patients received Mebendazole (NCT02465060). | _ |
| Escitalopram oxalate* | Anti-depressant | 2002 | Under phase trial | 8 | Pre-clinical | Terminated due to more adverse events in placebo group | _ |
| Verapamil | Calcium channel blocker | 1998 | Under phase trial | 9, 12, 38, 43 | Pre-clinical, Prospective study | No recent randomized clinical trial conducted. | _ |
| Vancomycin | Antibiotic | 1958 | Under phase trial | 4, 5 | Pre-clinical | No study results published | _ |
| Sildenafil | Erectile dysfunction | 1998 | Under phase trial | 31, | Pre-clinical | Results not yet published. | _ |
| Pirfenidone | Idiopathic pulmonary fibrosis | 2014 | Under phase trial | 9, 12 | Pre-clinical | NCT03177291 - Active, not recruiting; first posted 2017. Currently closed to accrual | _ |
| Digoxin | Arrhythmias | 2002 | Under phase trial | 1, 2, 42 | Pre-clinical | Terminated (NCT00281021). Digoxin did not increase the response rate of erlotinib in the treatment of progressive NSCLC. | Abstract |
| Auranofin | Swollen joints and morning stiffness | 1985 | Under phase trial | 12, 28 | Pre-clinical  Retrospective | Withdrawn (NCT02126527 - no participants enrolled). NCT01737502 - Recruiting | _ |
| Risperidone | Bipolar, schizophrenia | 1993 | Under phase trial | 30 | Pre-clinical | Pharmacokinetic analysis completed in adenocarcinoma patients including LC | - |
| Ibuprofen | Anti-inflammatory and antipyretic |  | Under phase trial | 9 | Pre-clinical | Study in planning stage, no published results. | - |
| Canakinumab | Juvenile idiopathic arthritis | 2009 | Under phase trial | 3,4 | Pre-clinical | Trial protocol published; results are not available yet. | - |
| Cannabidiol | Anti-seizure | 2018 | Under phase trial | 27 | Pre-clinical | Withdrawn (NCT02675842 - funding) | _ |
| Loratadine | Hay fever | 2002 | Under phase trial | 26 | Pre-clinical | No study available in LC patients with loratadine, there is a study on desloratadine NCT02646020.  Improved survival reported in meta-analysis. | ([Ildikó Fritz](https://pubmed.ncbi.nlm.nih.gov/?term=Fritz%20I%5BAuthor%5D) et al. 2021 Meta-analysis) |
| beta-blockers**^a^** | Beta-adrenoceptors inhibitor | _ | Under phase trial | 34 | Retrospective and Pre-clinical | Findings do not support an overall survival advantage in patients with lung cancer using beta-blocker therapy. | Coelho et al. 2020 |
| Statins**^a^** | HMG-CoA reductase inhibitors | 1987 | Under phase trial | 1, 4, 6, 8 | Pre-clinical  Retrospective | Beneficial effect of mortality and overall survival for patients with early-stage NSCLC in retrospective studies analyzed. In this meta-analysis the drug showed no effect in RCTs. | [Dao-Kui Xia](https://pubmed.ncbi.nlm.nih.gov/?term=Xia%20DK%5BAuthor%5D) et al 2019 |

**Supplementary table S2:** List of studies with no study results to further evaluate efficacy or safety in lung cancer patients for 24 non-chemo drugs. (*) represents the drugs found in clinicaltrials.gov upon search with terms such as selective serotonin reuptake inhibitors, beta-blockers, calcium channel blockers. (^a^) represents unspecific non-chemo drug category with major evidence from meta-analysis.

| **No.** | **All Non-Cancer drugs** | **NCT#** |
| --- | --- | --- |
| 1 | Lovastatin | no study found |
| 2 | Lopinavir | no study found |
| 3 | Sertraline | no study found |
| 4 | Tigecycline | no study found |
| 5 | Ritonavir | no study found |
| 6 | Mefloquine | no study found |
| 7 | Mepacrine | no study found |
| 8 | Niclosamide | no study found |
| 9 | Tanespimycin | no study found |
| 10 | Spiperone | no study found |
| 11 | Clomipramine | no study found |
| 12 | Prazosin | no study found |
| 13 | Pyrvinium Pamoate | no study found |
| 14 | Prenylamine | no study found |
| 15 | Anisomycin | no study found |
| 16 | Trichostatin A | no study found |
| 17 | Monensin | no study found |
| 18 | Cloperastine | no study found |
| 19 | Securinine | no study found |
| 20 | Fingolimod | no study found |
| 21 | Loratadine | no study found |
| 22 | Pitavastatin | no study found |
| 23 | Doxazosin | no study found |

**Supplementary Table S3:** List of all 23 non-chemo drugs identified from 45 literatures excluding drugs suggested from preclinical studies or reviews.

| **No.** | **All Non-Cancer drugs** | **NCT# for Lung Cancer with (http)** | **count** |
| --- | --- | --- | --- |
| 1 | Nelfinavir | NCT00589056, NCT01447589, NCT01108666, NCT00791336, NCT03050060  (https://classic.clinicaltrials.gov/ct2/results?cond=Lung+Cancer&term=Nelfinavir&cntry=&state=&city=&dist=) | 5 |
| 2 | Minocycline | NCT01636934, NCT01317550, NCT01048983, NCT00473083  (https://classic.clinicaltrials.gov/ct2/results?cond=Lung+Cancer&term=Minocycline&cntry=&state=&city=&dist=) | 4 |
| 3 | Atovaquone | NCT04648033, NCT02628080  (https://classic.clinicaltrials.gov/ct2/results?cond=Lung+Cancer&term=Atovaquone&cntry=&state=&city=&dist=) | 2 |
| 4 | Vancomycin | NCT03546829, NCT05777603, NCT05354102  (https://classic.clinicaltrials.gov/ct2/results?cond=Lung+Cancer&term=Vancomycin&cntry=&state=&city=&dist=) | 3 |
| 5 | Rosuvastatin | NCT02317016, NCT05926180, NCT00966472  (https://classic.clinicaltrials.gov/ct2/results?cond=Lung+Cancer&term=Rosuvastatin&cntry=&state=&city=&dist=) | 3 |
| 6 | Apricoxib | NCT01532362, NCT00771953, NCT00652340  (https://classic.clinicaltrials.gov/ct2/results?cond=Lung+Cancer&term=Apricoxib&cntry=&state=&city=&dist=) | 3 |
| 7 | Hydroxychloroquine | NCT00728845, NCT00809237, NCT04735068, NCT05647330, NCT01649947, NCT02722369, NCT01026844, NCT00977470, NCT02470468, NCT05733000  (https://classic.clinicaltrials.gov/ct2/results?cond=Lung+Cancer&term=Hydroxychloroquine&cntry=&state=&city=&dist=) | 10 |
| 8 | Pirfenidone | NCT05801133, NCT03177291, NCT04467723, NCT05704166  (https://classic.clinicaltrials.gov/ct2/results?cond=Lung+Cancer&term=Pirfenidone&cntry=&state=&city=&dist=) | 4 |
| 9 | Disulfiram | NCT00312819  (https://classic.clinicaltrials.gov/ct2/results?cond=Lung+Cancer&term=Disulfiram&cntry=&state=&city=&dist=) | 1 |
| 10 | Clarithromycin | NCT02416570, NCT02852083 (https://classic.clinicaltrials.gov/ct2/results?cond=Lung+Cancer&term=Clarithromycin&cntry=&state=&city=&dist=) | 2 |
| 11 | Bisphosphonates  Risedronate  (Found when searched for CCB) | NCT00765687, NCT02622607, NCT01004510, NCT04325776, NCT00264420, NCT03669523, NCT00896454, NCT00172042, NCT01204203, NCT05614518  (https://classic.clinicaltrials.gov/ct2/results?cond=Lung+Cancer&term=Bisphosphonates&cntry=&state=&city=&dist=)  NCT03861091  (https://classic.clinicaltrials.gov/ct2/results?cond=Lung+Cancer&term=Risedronate&cntry=&state=&city=&dist=) | 11 |
| 12 | Simvastatin | NCT00452244, NCT00452634, NCT01441349, NCT04985201, NCT04698941, NCT01156545, NCT02197234  (https://classic.clinicaltrials.gov/ct2/results?cond=Lung+Cancer&term=Simvastatin&cntry=&state=&city=&dist=) | 7 |
| 13 | Celecoxib | NCT00020878, NCT00030407, NCT00055978, NCT00030420, NCT01503385, NCT00046839, NCT00346801, NCT00062179, NCT00068653, NCT00527982, NCT00104767, NCT00520845, NCT00072072, NCT00653250, NCT00073866, NCT00300729, NCT00274898, NCT00088959, NCT00108186, NCT00062101, NCT00058006, NCT00499655, NCT00070486, NCT01041781, NCT00211952, NCT00181532, NCT00037817, NCT01258868, NCT00442754, NCT01143545, NCT01313429, NCT03710876, NCT02054104.  (https://classic.clinicaltrials.gov/ct2/results?cond=Lung+Cancer&term=Celecoxib&cntry=&state=&city=&dist=) | 33 |
| 14 | Rofecoxib | NCT00385606 (https://classic.clinicaltrials.gov/ct2/results?cond=Lung+Cancer&term=Rofecoxib&cntry=&state=&city=&dist=) | 1 |
| 15 | Auranofin | NCT02126527, NCT01737502  (https://classic.clinicaltrials.gov/ct2/results?cond=Lung+Cancer&term=Auranofin&cntry=&state=&city=&dist=) | 2 |
| 16 | Nitroglycerin | NCT01210378, NCT00616031, NCT01171170, NCT00886405, NCT04338867, NCT05172739.  (https://classic.clinicaltrials.gov/ct2/results?cond=Lung+Cancer&term=Nitroglycerin&cntry=&state=&city=&dist=) | 6 |
| 17 | Mebendazole | NCT04771520, NCT04870034, NCT02465060  (https://classic.clinicaltrials.gov/ct2/results?cond=Lung+Cancer&term=Mebendazole&cntry=&state=&city=&dist=) | 3 |
| 18 | Itraconazole | NCT03664115, NCT02357836, NCT00769600, NCT02157883, NCT01752023, NCT00585195, NCT02349633, NCT05150691  (https://classic.clinicaltrials.gov/ct2/results?cond=Lung+Cancer&term=Itraconazole&cntry=&state=&city=&dist=) | 8 |
| 19 | Sodium valproate | NCT01203735, NCT00084981, NCT00759824, NCT00996060, NCT00634205  (https://classic.clinicaltrials.gov/ct2/results?cond=Lung+Cancer&term=Sodium+valproate&cntry=&state=&city=&dist=) | 5 |
| 20 | Propranolol | NCT05979818, NCT00527319  (https://classic.clinicaltrials.gov/ct2/results?cond=Lung+Cancer&term=Propranolol&cntry=&state=&city=&dist=) | 2 |
| 21 | Etodolac | NCT00527319 (https://classic.clinicaltrials.gov/ct2/results?cond=Lung+Cancer&term=Etodolac&cntry=&state=&city=&dist=) | 1 |
| 22 | Atorvastatin | NCT01259284  (https://classic.clinicaltrials.gov/ct2/results?cond=Lung+Cancer&term=Atorvastatin&cntry=&state=&city=&dist=) | 1 |
| 23 | Metformin | NCT02115464, NCT02285855, NCT05445791, NCT02019979, NCT03086733, NCT01717482, NCT01997775, NCT02109549, NCT04931017, NCT03874000, NCT03048500, NCT03994744, NCT04001725, NCT03071705, NCT02186847, NCT04170959, NCT01864681, NCT00659568, NCT02145559, NCT02431676, NCT01578551, NCT03709147  (https://classic.clinicaltrials.gov/ct2/results?cond=Lung+Cancer&term=Metformin&cntry=&state=&city=&dist=) | 22 |
| 24 | Quinacrine | NCT01839955  (https://classic.clinicaltrials.gov/ct2/results?cond=Lung+Cancer&term=Quinacrine&cntry=&state=&city=&dist=) | 1 |
| 25 | Romidepsin | NCT00086827, NCT01302808, NCT00020202, NCT00037817, NCT00094978, NCT01638533  (https://classic.clinicaltrials.gov/ct2/results?cond=Lung+Cancer&term=Romidepsin&cntry=&state=&city=&dist=) | 6 |
| 26 | Aspirin/Acetylsalicylic acid | NCT01352962, NCT01727427, NCT02169271, NCT01058902, NCT03532698, NCT06018688, NCT03543683, NCT04184921, NCT01707823, NCT02123849, NCT05988697, NCT02387086, NCT03654105  (https://classic.clinicaltrials.gov/ct2/results?cond=Lung+Cancer&term=Aspirin&cntry=&state=&city=&dist=) | 13 |
| 27 | Mirtazapine | NCT04748523, NCT02650544, NCT04155008, NCT01216371  (https://classic.clinicaltrials.gov/ct2/results?cond=Lung+Cancer&term=Mirtazapine&cntry=&state=&city=&dist=) | 4 |
| 28 | Risperidone | NCT02875340  (https://classic.clinicaltrials.gov/ct2/results?cond=Lung+Cancer&term=Risperidone&cntry=&state=&city=&dist=) | 1 |
| 29 | Zoledronic acid | NCT02622607, NCT00172042, NCT01004510, NCT04325776, NCT02480634, NCT00559897, NCT01737216, NCT00762346, NCT00265200, NCT00099541, NCT00264420, NCT00365105, NCT03958565, NCT00086268, NCT00765687, NCT00003884, NCT00874211, NCT01204203, NCT01951586  (https://classic.clinicaltrials.gov/ct2/results?cond=Lung+Cancer&term=Zoledronic+acid&cntry=&state=&city=&dist=) | 19 |
| 30 | Aprepitant | NCT02364804, NCT02445872, NCT02646020, NCT02161991, NCT00588835, NCT03571126, NCT02407600  (https://classic.clinicaltrials.gov/ct2/results?cond=Lung+Cancer&term=Aprepitant&cntry=&state=&city=&dist=) | 7 |
| 31 | Eprenetapopt | NCT04383938 (https://classic.clinicaltrials.gov/ct2/results?cond=Lung+Cancer&term=Eprenetapopt&cntry=&state=&city=&dist=) | 1 |
| 32 | cannabidiol | NCT02675842, NCT03763851  (https://classic.clinicaltrials.gov/ct2/results?cond=Lung+Cancer&term=cannabidiol&cntry=&state=&city=&dist=) | 2 |
| 33 | Ibuprofen | NCT04907864, NCT05834569  (https://classic.clinicaltrials.gov/ct2/results?cond=Lung+Cancer&term=Ibuprofen&cntry=&state=&city=&dist=) | 2 |
| 34 | Digoxin | NCT05926180, NCT00281021  (https://classic.clinicaltrials.gov/ct2/results?cond=Lung+Cancer&term=Digoxin&cntry=&state=&city=&dist=) | 2 |
| 35 | Retinoic acid | NCT01041833, NCT00002586, NCT01048645, NCT04919369, NCT00617409, NCT02712905 (8 Trials on All trans retinoic acid and tretinoin excluded)  (https://classic.clinicaltrials.gov/ct2/results?cond=Lung+Cancer&term=Retinoic+acid&cntry=&state=&city=&dist=) | 6 |
| 36 | Nortriptyline | NCT02881125  (https://classic.clinicaltrials.gov/ct2/results?cond=Lung+Cancer&term=Nortriptyline&cntry=&state=&city=&dist=) | 1 |
| 37 | Pravastatin | NCT00433498  (https://classic.clinicaltrials.gov/ct2/results?cond=Lung+Cancer&term=Pravastatin&cntry=&state=&city=&dist=) | 1 |
| 38 | Thalidomide | NCT00061919, NCT00053300, NCT00004859, NCT00114192, NCT00281827, NCT00025285, NCT04382300, NCT02778893, NCT03341494, NCT02387086, NCT00027638, NCT02963610, NCT00049296, NCT03062800  (https://classic.clinicaltrials.gov/ct2/results?cond=Lung+Cancer&term=Thalidomide&cntry=&state=&city=&dist=) | 14 |
| 39 | Anastrozole | NCT00932152, NCT02779751 (https://classic.clinicaltrials.gov/ct2/results?cond=Lung+Cancer&term=Anastrozole&cntry=&state=&city=&dist=) | 2 |
| 40 | Naproxen | NCT01612975 (https://classic.clinicaltrials.gov/ct2/results?cond=Lung+Cancer&term=Naproxen&cntry=&state=&city=&dist=) | 1 |
| 41 | Canakinumab | NCT05725343, NCT04905316, NCT03447769, NCT03968419, NCT03631199, NCT04789681, NCT03626545, NCT06038526, NCT03064854, NCT02900664 (https://classic.clinicaltrials.gov/ct2/results?cond=Lung+Cancer&term=Canakinumab&cntry=&state=&city=&dist=) | 10 |
| 42 | Statins | NCT05636592, NCT01169051 (https://classic.clinicaltrials.gov/ct2/results?cond=Lung+Cancer&term=Statins&cntry=&state=&city=&dist=) | 2 |
| 43 | Pioglitazone | NCT00780234, NCT00923949, NCT05919147, NCT01342770, NCT02852083 (https://classic.clinicaltrials.gov/ct2/results?cond=Lung+Cancer&term=Pioglitazone&cntry=&state=&city=&dist=) | 5 |
| 44 | Enalapril maleate | NCT01754909 (https://classic.clinicaltrials.gov/ct2/results?cond=Lung+Cancer&term=Enalapril+maleate&cntry=&state=&city=&dist=) | 1 |
| 45 | Perindopril | NCT04980716 (https://classic.clinicaltrials.gov/ct2/results?cond=Lung+Cancer&term=Perindopril&cntry=&state=&city=&dist=) | 1 |
| 46 | Metoprolol | NCT01281787, NCT04980716 (https://classic.clinicaltrials.gov/ct2/results?cond=Lung+Cancer&term=Metoprolol&cntry=&state=&city=&dist=) | 2 |
| 47 | Spironolactone | NCT04980716 (https://classic.clinicaltrials.gov/ct2/results?cond=Lung+Cancer&term=Spironolactone&cntry=&state=&city=&dist=) | 1 |
| 48 | Trimetazidine hydrochloride | NCT04980716 (https://classic.clinicaltrials.gov/ct2/results?cond=Lung+Cancer&term=Trimetazidine+hydrochloride&cntry=&state=&city=&dist=) | 1 |
| 49 | Sildenafil | NCT00752115, NCT02349633 (https://classic.clinicaltrials.gov/ct2/results?cond=Lung+Cancer&term=Sildenafil&cntry=&state=&city=&dist=) | 2 |
| 50 | Arsenic trioxide | NCT02066870, NCT00075426, NCT01470248 (https://classic.clinicaltrials.gov/ct2/results?cond=Lung+Cancer&term=Arsenic+trioxide&cntry=&state=&city=&dist=) | 3 |
| 51 | Tocotrienol | NCT02644252 (https://classic.clinicaltrials.gov/ct2/results?cond=Lung+Cancer&term=Tocotrienol&cntry=&state=&city=&dist=) | 1 |
| 52 | Doxycycline | NCT01411202, NCT00531934, NCT00803842, NCT01465802 (https://classic.clinicaltrials.gov/ct2/results?cond=Lung+Cancer&term=Doxycycline&cntry=&state=&city=&dist=) | 4 |
| 53 | Desipramine | NCT01719861 (https://classic.clinicaltrials.gov/ct2/results?cond=Lung+Cancer&term=Desipramine&cntry=&state=&city=&dist=) | 1 |
| 54 | Sirolimus  Rapamycin | NCT00375245, NCT05997056, NCT01522820, NCT02145559, NCT00923273, NCT03217669, NCT05840510, NCT00993499, NCT01737502, NCT01383668, NCT04348292, NCT02126527, NCT00098462 (https://classic.clinicaltrials.gov/ct2/results?cond=Lung+Cancer&term=Sirolimus&cntry=&state=&city=&dist=)  NCT00555256, NCT00993499, NCT01390818, NCT03190174, NCT00375245 (https://classic.clinicaltrials.gov/ct2/results?cond=Lung+Cancer&term=Rapamycin&cntry=&state=&city=&dist=) | 18 |
| 55 | Tilarginine | NCT03236935 (https://classic.clinicaltrials.gov/ct2/results?cond=Lung+Cancer&term=Tilarginine&cntry=&state=&city=&dist=) | 1 |
| 56 | Denosumab | NCT02129699, NCT03669523, NCT03958565, NCT00896454, NCT01951586 (https://classic.clinicaltrials.gov/ct2/results?cond=Lung+Cancer&term=Denosumab&cntry=&state=&city=&dist=) | 4 |
| 57 | calcium channel blockers (CCB)  carboxyamidotriazole | NCT00003869, NCT00019019 (https://classic.clinicaltrials.gov/ct2/results?cond=Lung+Cancer&term=calcium+channel+blockers&cntry=&state=&city=&dist=) | 2 |
| 58 | Selective serotonin reuptake inhibitors  fluoxetine  Olanzapine  escitalopram oxalate | NCT00005850,  NCT03571126,    NCT00387348  (https://classic.clinicaltrials.gov/ct2/results?cond=Lung+Cancer&term=selective+serotonin+reuptake+inhibitors&cntry=&state=&city=&dist=) | 3 |

**Supplementary table S4:** non-chemo medications (n=61) are marked in (blue). NCT# with recruitment status completed or completed (has results) or unknown status are marked as (green), RCT# with recruiting, not yet recruiting, enrolling, active not recruiting, terminated, suspended, withdrawn are marked as (red). Trial status for drugs with recruiting, not yet recruiting, enrolling, active not recruiting, terminated, suspended, withdrawn and finally with no published pivotal research are marked as (purple).

| **Drugs (Clinical Phase)** | **Trial Identifier** | **Drugs** | **Clinical findings** | **Reference** |
| --- | --- | --- | --- | --- |
| Arsenic trioxide Phase-II) | NCT01470248 | Post first-line chemo arsenic trioxide infusion | In 12% patients had stable disease and 88% had progressive disease. | [1] |
| Nelfinavir (Phase-II) | NCT00589056 | Nelfinavir Concurrently with CT-RT | ORR was achieved in 94% locally advanced NSCLC patients with no dose limiting toxicities. The median PFS 11.7 months and OS of 41.1 months was noted. | [2] |
| Valproic acid (Phase-II) | NCT00634205  NCT00759824 | Post-failure of platinum-based chemotherapy treatment with doxorubicin plus valproic acid  Doxorubicin, Vindesine, and cyclophosphamide along with valproic acid | Disease control rate was in 36% patients and 16% with partial response against malignant mesothelioma. No drug related deaths were reported.  The primary objective of 6 months PFS was achieved in 6% patients, and 88% patients had severe hematological grade 2 adverse events. | [3]  [4] |
| Romidepsin (Phase-II) | NCT00086827  (1^st^ Registered in 2004) | Post platinum-based chemo (69% with RT), Romidepsin infusion/week | Poor objective response rate found in SCLC patients, 19% had desired response and 25% had severe grade 3 or 4 adverse events. | [5] |
| Desipramine (Phase-II) | NCT01719861 | Post 1^st^ line chemotherapy failure desipramine/day | 67% patients were diagnosed with SCLC and NET of pulmonary origin. Very poor PFS and OS was noted with treatment related adverse events. | [6] |
| Hydroxychloroquine (Phase II) | NCT01649947 | carboplatin, paclitaxel, bevacizumab and hydroxychloroquine | Clinical benefit was 53% with median 3.3 months of median PFS. In 9 KRAS positive patients 44% ORR and median PFS was 6.4 months. | [7] |

**Supplementary Table S5:** Phase II open label non-randomized single-arm clinical studies on lung cancer patients with chemotherapy and 6 non-chemo drugs are presented. OS; Overall Survival, PFS; Progression Free Survival, ORR; Overall response rate, NSCLC; Non-small cell Lung Cancer, SCLC; Small Cell Lung Cancer, NET; Neuroendocrine tumor, KRAS; Kirsten rat sarcoma viral oncogene homolog.

**References: 1 study included in conclusion**

| 1 | Indomethacin | no study found |
| --- | --- | --- |
| 2 | Levamisole | no study found |
| 3 | Halimide | no study found |
| 4 | Dihydroartemisinic | no study found |
| 5 | Carglumic acid | no study found |
| 6 | Fluphenazine | no study found |
| 7 | Pimozide | no study found |
| 8 | Trifluoperazine | no study found |
| 9 | Perphenazine | no study found |
| 10 | Clobetasol propionate | no study found |
| 11 | Albendazole | no study found |
| 12 | Bezafibrate | no study found |
| 13 | Potassium Antimonyl Tartrate | no study found |
| 14 | Rifabutin | no study found |
| 15 | Ouabain | no study found |
| 16 | Meclofenamate | no study found |
| 17 | Miconazole | no study found |
| 18 | Thioridazine | no study found |
| 19 | Menadione | no study found |
| 20 | Promazine | no study found |
| 27 | Vidarabine | no study found |
| 28 | Clioquinol | no study found |
| 29 | Ellipticine | no study found |
| 30 | Cantharidin | no study found |
| 31 | Puromycin | no study found |
| 32 | Darapladib | no study found |
| 33 | Rilapladib | no study found |
| 34 | Tilarginine | No study found |
| 35 | Digitoxin | no study found |
| 36 | Isoprenaline | no study found |
| 37 | Fumonisin B1 | no study found |
| 38 | Indisulam | no study found |
| 39 | Sotrastaurin | no study found |
| 40 | Artemisinin | no study found |
| 41 | fluspirilene | no study found |
| 42 | Penfluridol | no study found |
| 43 | Sulfasalazine | no study found |
| 44 | Mefloquine | no study found |
| 45 | Clemastine | no study found |
| 46 | Sulindac Acid | no study found |
| 47 | Amino salicylic Acid | no study found |
| 48 | Ibudilast | no study found |
| 49 | Anakinra | no study found |
| 50 | Rilonacept | no study found |
| 51 | Panzem | no study found |
| 52 | Ezetimibe | no study found |
| 53 | Deptropine | no study found |
| 54 | Azacyclonol | no study found |
| 55 | Verapamil | no study found |
| 56 | Pantoprazole | no study found |
| 57 | Tegecycline | no study found |

**Supplementary table S6:** Drugs with no available studies these drugs were suggested for human use based on pre-clinical evidence. Here is the evidence that drugs reported are not in pre-clinical or in clinical phase also not registered in ClinicalTrials.gov

| **Drug name** | **NCT#** | **Design Bias (1-3)**  **A B C** | | | **Selection Bias (1-3)** | **Reporting Bias (1-3)** | **Overall**  **Score (1-9)** |
| --- | --- | --- | --- | --- | --- | --- | --- |
| Disulfiram | NCT00312819 | * | * | * | *** | ** | 8 |
| Metformin | NCT01864681 | * | * | * | *** | *** | 9 |
| Minocycline | NCT00473083 | * | - | * | *** | ** | 7 |
|  | [NCT01636934](http://clinicaltrials.gov/show/NCT01636934) | * | * | * | *** | *** | 9 |
| Pravastatin | NCT00433498 | * | * | * | *** | *** | 9 |
| Aprepitant | NCT02161991 | * | * | * | *** | *** | 9 |
| Aspirin | NCT02123849 | * | * | - | ** | ** | 6 |
|  | NCT02169271 | * | * | * | *** | ** | 8 |
| Zoledronic acid  Denosumab | NCT00172042 | * | - | * | ** | ** | 5 |
|  | NCT00264420 | * | * | * | *** | *** | 9 |
|  | NCT01951586  NCT02129699 | * | - | * | *** | ** | 7 |
| Thalidomide | NCT00004859 | * | - | * | *** | ** | 7 |
|  | NCT00061919 | * | * | * | *** | *** | 9 |
| Rofecoxib  Celecoxib  Apricoxib | NCT00385606 | * | - | * | *** | ** | 7 |
|  | NCT01041781 | * | * | * | *** | *** | 9 |
|  | NCT00300729 | * | * | * | *** | *** | 9 |
|  | NCT00652340 | * | - | * | *** | ** | 7 |
| Carboxyaminoimidazole | NCT00003869 | * | * | * | *** | *** | 9 |
| Itraconazole | NCT03664115 | * | - | * | *** | ** | 7 |
| Risedronate | NCT03861091 | * | * | * | ** | * | 6 |
| 13-cis Retinoic acid | NCT00002586 | * | * | * | *** | *** | 9 |
| Etodolac plus propranolol (VT-122) | NCT00527319 | * | - | * | ** | ** | 6 |
| Pioglitazone and clarithromycin | NCT02852083 | * | - | * | *** | * | 6 |
| Pioglitazone | NCT00780234 | * | * | * | *** | *** | 9 |
| Nitroglycerine | NCT01171170 | * | - | * | *** | ** | 7 |
|  | NCT04338867 | * | - | * | ** | ** | 6 |
| Doxycycline | NCT00531934 | * | - | * | *** | ** | 7 |
|  | NCT01465802 | * | - | * | *** | ** | 7 |
| Simvastatin | NCT00452244 | * | - | * | *** | ** | 7 |
|  | NCT01156545 | * | - | * | *** | ** | 7 |

**Supplementary Table S7:** Each study included for final assessment is graded for biases based on study design; (A) randomized (*), (B) open (-) blinded (*), (C) placebo-controlled (*); selection bias (low - **/** ***), (moderate - **/** **), (high - **/** *); reporting bias (low - **/** ***), (moderate - **/** **), (high - **/** *). Based on the weightage of (*) overall bias score (1-9) was assessed, score of 5 or > is moderate-high grade literatures were included.

**Supplementary Table S8:** List of Articles included or excluded; Authors marked as Green are the articles included in final analysis, Articles common to both clinicaltrials.gov and Google scholar are highlighted in red.

| Author | Article title Google Scholar | Article title ClinicalTrials.Gov | Author |
| --- | --- | --- | --- |
| Curtis Tilves | A behavioral weight-loss intervention, but not metformin, decreases a marker of gut barrier permeability: results from the SPIRIT randomized trial | A Multicenter Double-blind Phase II Study of Metformin With Gefitinib as First-line Therapy of Locally Advanced NoneSmall-cell Lung Cancer | Kun-lin Li |
| Kun-lin Li | A Multicenter Double-blind Phase II Study of Metformin With Gefitinib as First-line Therapy of Locally Advanced NoneSmall-cell Lung Cancer | A Phase IIb Trial Assessing the Addition of Disulfiram to Chemotherapy for the Treatment of Metastatic Non-Small Cell Lung Cancer | HOVAV NECHUSHTAN |
| Byoung Chul Cho | A phase 1b/2 study of PF-06747775 as monotherapy or in combination with Palbociclib in patients with epidermal growth factor receptor mutant advanced non-small cell lung cancer | A phase I study of high-dose rosuvastatin with standard dose erlotinib in patients with advanced solid malignancies | Glenwood D. Goss |
| Glenwood D. Goss | A phase I study of high-dose rosuvastatin with standard dose erlotinib in patients with advanced solid malignancies | A Phase I Trial to Determine the Optimal Biological Dose of Celecoxib when Combined with Erlotinib in Advanced Non-Small Cell Lung Cancer | Karen L. Reckamp |
| M. E. Lacouture | A phase II study (ARCHER 1042) to evaluate prophylactic treatment of dacomitinib-induced dermatologic and gastrointestinal adverse events in advanced non-smallcell lung cancer | A Randomized Phase II Chemoprevention Trial of 13-CIS Retinoic Acid with Or without α Tocopherol or Observation in Subjects at High Risk for Lung Cancer | Karen Kelly |
| Jonathan W. Riess | A PHASE IIA STUDY REPOSITIONING DESIPRAMINE IN SMALL CELL LUNG CANCER AND OTHER HIGH-GRADE NEUROENDOCRINE TUMORS | A Randomized Phase II Study of Gefitinib Plus Simvastatin Versus Gefitinib Alone in Previously Treated Patients with Advanced Non–Small Cell Lung Cancer | Ji-Youn Han |
| HOVAV NECHUSHTAN | A Phase IIb Trial Assessing the Addition of Disulfiram to Chemotherapy for the Treatment of Metastatic Non-Small Cell Lung Cancer | A Randomized Phase II Study of Metformin plus Paclitaxel/Carboplatin/Bevacizumab in Patients with Chemotherapy-Naive Advanced or Metastatic Nonsquamous Non-Small Cell Lung Cancer | KRISTEN A. MARRONE |
| Muhammad Omer Jamil | A pilot study of zoledronic acid in the treatment of patients with advanced malignant pleural mesothelioma | Addition of Metformin to Concurrent Chemoradiation in Patients With Locally Advanced Non–Small Cell Lung Cancer The NRG-LU001 Phase 2 Randomized Clinical Trial | Heath Skinner |
| KRISTEN A. MARRONE | A Randomized Phase II Study of Metformin plus Paclitaxel/Carboplatin/Bevacizumab in Patients with Chemotherapy-Naive Advanced or Metastatic Nonsquamous Non-Small Cell Lung Cancer | Anti-angiogenic Therapy Using Thalidomide Combined With Chemotherapy in Small Cell Lung Cancer: A Randomized, Double-Blind, Placebo-Controlled Trial | Siow Ming Lee |
| Dingemans | A randomized phase II study comparing paclitaxel–carboplatin–bevacizumab with or without nitroglycerin patches in patients with stage IV non-squamous non-small-cell lung cancer: NVALT12 (NCT01171170)† | Aprepitant plus palonosetron and dexamethasone for prevention of chemotherapy-induced nausea and vomiting in patients receiving multiple-day cisplatin chemotherapy | H. F. Gao |
| Heudobler | A Randomized Phase II Trial Comparing the Efficacy and Safety of Pioglitazone, Clarithromycin and Metronomic Low-Dose Chemotherapy with Single-Agent Nivolumab Therapy in Patients with Advanced Non-small Cell Lung Cancer Treated in Second or Further Line (ModuLung) | Aprepitant triple therapy for the prevention of chemotherapy-induced nausea and vomiting following high-dose cisplatin in Chinese patients: a randomized, double-blind, placebo-controlled phase III trial | Zhihuang Hu |
| Robert L. Keith | A Randomized Phase II Trial of Pioglitazone for Lung Cancer Chemoprevention in High-Risk Current and Former Smokers | Canakinumab with and without pembrolizumab in patients with resectable non-small-cell lung cancer: CANOPY-N study design | Pilar Garrido |
| Barbara J. Gitlitz | A Randomized, Placebo-Controlled, Multicenter, Biomarker-Selected, Phase 2 Study of Apricoxib in Combination with Erlotinib in Patients with Advanced Non–Small-Cell Lung Cancer | Comparison of aprepitant versus desloratadine for EGFR‐TKI‐induced pruritus: A randomized phase 2 clinical trial | Ting Zhou |
| Siow Ming Lee | Anti-angiogenic Therapy Using Thalidomide Combined With Chemotherapy in Small Cell Lung Cancer: A Randomized, Double-Blind, Placebo Controlled Trial | Decreased Risk of Radiation Pneumonitis With Incidental Concurrent Use of Angiotensin-Converting Enzyme Inhibitors and Thoracic Radiation Therapy | Jordan Kharofa |
| Anne-Sophie Hamy | Celecoxib With Neoadjuvant Chemotherapy for Breast Cancer Might Worsen Outcomes Differentially by COX-2 Expression and ER Status: Exploratory Analysis of the REMAGUS02 Trial | Effect of celecoxib on survival in patients with advanced non-small cell lung cancer: A double blind randomised clinical phase III trial (CYCLUS study) by the Swedish Lung Cancer Study Group | Andrea Koch |
| Lotte Engell-Noerregaard | Clinical and Immunological Effects in Patients with Advanced Non-Small Cell Lung-Cancer after Vaccination with Dendritic Cells Exposed to an Allogeneic Tumor Cell Lysate | Effect of Metformin Plus Tyrosine Kinase Inhibitors Compared With Tyrosine Kinase Inhibitors Alone in Patients With Epidermal Growth Factor Receptor–Mutated Lung Adenocarcinoma A Phase 2 Randomized Clinical Trial | Oscar Arrieta |
| Ramesh Rengan | Clinical Outcomes of the HIV Protease Inhibitor Nelfinavir With Concurrent Chemoradiotherapy for Unresectable Stage IIIA/IIIB Non–Small Cell Lung Cancer A Phase 1/2 Trial | Effect of multiple-dose osimertinib on the pharmacokinetics of simvastatin and rosuvastatin | R. Donald Harvey |
| Suzanne J. Dilly | Clinical Pharmacokinetics of a Lipid‑Based Formulation of Risperidone, VAL401: Analysis of a Single Dose in an Open‑Label Trial of Late‑Stage Cancer Patients | Effects of statin exposure and lung cancer survival: A meta-analysis of observational studies | Yafei Chen |
| Ting Zhou | Comparison of aprepitant versus desloratadine for EGFR‐TKI‐induced pruritus: A randomized phase 2 clinical trial | Efficacy of metformin in combination with immune checkpoint inhibitors (anti-PD-1/anti-CTLA-4) in metastatic malignant melanoma | Muhammad Zubair Afzal |
| David E. Gerber | Concentration-dependent Early Antivascular and Antitumor Effects of Itraconazole in Non–Small Cell Lung Cancer | Factorial phase III randomised trial of rofecoxib and prolonged constant infusion of gemcitabine in advanced non-small-cell lung cancer: the GEmcitabine-COxib in NSCLC (GECO) study | Cesare Gridelli |
| Mimi I Hu | Denosumab for Treatment of Hypercalcemia of Malignancy in Patients with Solid Tumors or Hematological Malignancies Refractory to IV Bisphosphonates: A Single-Arm Multicenter Study | Investigation of atovaquone-induced spatial changes in tumour hypoxia assessed by hypoxia PET/CT in non-small cell lung cancer patients | Pauline Bourigault |
| Deplanque | Doxycycline for prevention of erlotinib-induced rash in patients with non-small-cell lung cancer (NSCLC) after failure of first-line chemotherapy: A randomized, open-label trial | Long-Term Efficacy and Safety of Zoledronic Acid in the Treatment of Skeletal Metastases in Patients with Nonsmall Cell Lung Carcinoma and Other Solid Tumors A Randomized, Phase III, Double-Blind, Placebo-Controlled Trial | Lee S. Rosen |
| Andrea Koch | Effect of celecoxib on survival in patients with advanced non-small cell lung cancer: A double blind randomised clinical phase III trial (CYCLUS study) by the Swedish Lung Cancer Study Group | Metabolic Responses to Metformin in Inoperable Early-stage Non–Small Cell Lung Cancer Treated With Stereotactic Radiotherapy Results of a Randomized Phase II Clinical Trial | Stephen G. Chun |
| Nan Bi | Effect of Concurrent Chemoradiation With Celecoxib vs Concurrent Chemoradiation Alone on Survival Among Patients With Non–Small Cell Lung Cancer With and Without Cyclooxygenase 2 Genetic Variants A Phase 2 Randomized Clinical Trial | Metformin in Combination With Chemoradiotherapy in Locally Advanced Non–Small Cell Lung Cancer The OCOG-ALMERA Randomized Clinical Trial | Theodoros Tsakiridis |
| Linda L. Garland | Effect of Intermittent Versus Continuous Low-Dose Aspirin on Nasal Epithelium Gene Expression in Current Smokers: A Randomized, Double-Blinded Trial | Metformin use and its effect on survival in diabetic patients with advanced non-small cell lung cancer | Oscar Arrieta |
| Cesare Gridelli | Factorial phase III randomised trial of rofecoxib and prolonged constant infusion of gemcitabine in advanced non-small-cell lung cancer: the GEmcitabine-COxib in NSCLC (GECO) study | Multicenter, randomized, phase 2 study of zoledronic acid in combination with docetaxel and carboplatin in patients with unresectable stage IIIB or stage IV non-small cell lung cancer | Kishan J. Pandya |
| Baptiste Pichon | High-Dose Hypofractionated Radiation Therapy for Noncompressive Vertebral Metastases in Combination With Zoledronate: A Phase 1 Study | Nitroglycerin Plus Whole Intracranial Radiation Therapy for Brain Metastases in Patients With Non-Small Cell Lung Cancer: A Randomized, Open-Label, Phase 2 Clinical Trial | Oscar Arrieta |
| Karthick Vishwanathan | Impact of Disease and Treatment Response in Drug–Drug Interaction Studies: Osimertinib and Simvastatin in Advanced Non-Small Cell Lung Cancer | Pan Canadian Rash Trial: A Randomized Phase III Trial Evaluating the Impact of a Prophylactic Skin Treatment Regimen on Epidermal Growth Factor Receptor-Tyrosine Kinase Inhibitor–Induced Skin Toxicities in Patients With Metastatic Lung Cancer | Barbara Melosky |
| Krista C.J. Wink | Improved progression free survival for patients with diabetes and locally advanced non-small cell lung cancer (NSCLC) using metformin during concurrent chemoradiotherapy | Patient-derived xenografts faithfully replicated clinical outcome in a phase II co-clinical trial of arsenic trioxide in relapsed small cell lung cancer | Taofeek K. Owonikoko |
| Rui Han | Low BMI patients with advanced EGFR mutation-positive NSCLC can get a better outcome from metformin plus EGFR-TKI as first-line therapy: A secondary analysis of a phase 2 randomized clinical trial | Phase I/II Trial of a COX-2 Inhibitor With Limited Field Radiation for Intermediate Prognosis Patients Who Have Locally Advanced Non–Small-Cell Lung Cancer: Radiation Therapy Oncology Group 0213 | Elizabeth Gore |
| Bernardo Bonanni | Low-Dose Aspirin in High-Risk Individuals With Screen-Detected Subsolid Lung Nodules: A Randomized Phase II Trial | Phase Ib study of eprenetapopt (APR-246) in combination with pembrolizumab in patients with advanced or metastatic solid tumors | H. Park |
| Jenny T. Mao | Lung Cancer Chemoprevention with Celecoxib in Former Smokers | Phase II Study of Docetaxel and Celecoxib, a Cyclooxygenase-2 Inhibitor, in Elderly or Poor Performance Status (PS2) Patients with Advanced Non-small Cell Lung Cancer | Shirish M. Gadgeel |
| Stephen G. Chun | Metabolic Responses to Metformin in Inoperable Early-stage Non–Small Cell Lung Cancer Treated With Stereotactic Radiotherapy Results of a Randomized Phase II Clinical Trial | Phase 2 Study of Pemetrexed and Itraconazole as Second-Line Therapy for Metastatic Non-Squamous Non-Small Cell Lung Cancer | Charles M. Rudin |
| Michael Skwarski | Mitochondrial Inhibitor Atovaquone Increases Tumor Oxygenation and Inhibits Hypoxic Gene Expression in Patients with Non-Small Cell Lung Cancer | Phase II trial of thalidomide with chemotherapy and as maintenance therapy for patients with poor prognosis small-cell lung cancer | Siow Ming Lee |
| Bart J.T. Reymen | Nitroglycerin as a radiosensitizer in non-small cell lung cancer: Results of a prospective imaging-based phase II trial | Phase III Randomized, Placebo-Controlled, Double-Blind Trial of Celecoxib in Addition to Standard Chemotherapy for Advanced Non–Small-Cell Lung Cancer With Cyclooxygenase-2 Overexpression: CALGB 30801 (Alliance) | Martin J. Edelman |
| Oscar Arrieta | Nitroglycerin Plus Whole Intracranial Radiation Therapy for Brain Metastases in Patients With Non-Small Cell Lung Cancer: A Randomized, Open-Label, Phase 2 Clinical Trial | Phase I and pharmacokinetic study of docetaxel, irinotecan, and celecoxib in patients with advanced non-small cell lung cancer | Athanassios Argiris |
| Muhammed Murtaza | Non-invasive analysis of acquired resistance to cancer therapy by sequencing of plasma DNA | Phase I trial of docetaxel and thalidomide: a regimen based on metronomic therapeutic principles | Sharon L. Sanborn |
| T. Tsakiridis | OCOG-ALMERA: A phase II trial investigating the ability of metformin to chemo-radio-sensitize and prevent recurrence in locally advanced (LA) non-small cell lung cancer (NSCLC) | PHASE II STUDY OF CELECOXIB AND DOCETAXEL IN NONSMALL CELL LUNG CANCER (NSCLC) PATIENTS WITH PROGRESSION AFTER PLATINUM-BASED THERAPY | Bryan J. Schneider |
| Taofeek K. Owonikoko | Patient-derived xenografts faithfully replicated clinical outcome in a phase II co-clinical trial of arsenic trioxide in relapsed small cell lung cancer | Pyrotinib combined with thalidomide in advanced non-small-cell lung cancer patients harboring HER2 exon 20 insertions (PRIDE): protocol of an open-label, singlearm phase II trial | Xinghao Ai |
| Michal Hensler | Peripheral gene signatures reveal distinct cancer patient immunotypes with therapeutic implications for autologous DC-based vaccines | Randomized, double-blind phase II study to compare nitroglycerin plus oral vinorelbine plus cisplatin with oral vinorelbine plus cisplatin alone in patients with stage IIIB/IV non-small cell lung cancer (NSCLC) | N. Reinmuth |
| Jack M. Su | Phase 1 Study of Valproic Acid in Pediatric Patients with Refractory Solid or CNS Tumors: A Children's Oncology Group Report | Randomized phase III trial to evaluate radiopharmaceuticals and zoledronic acid in the palliation of osteoblastic metastases from lung, breast, and prostate cancer. Report of the NRG Oncology RTOG 0517 trial | Michael J. Seider |
| Charles M. Rudin | Phase 2 Study of Pemetrexed and Itraconazole as Second-Line Therapy for Metastatic Nonsquamous Non–Small-Cell Lung Cancer | Randomized, Double-Blind, Placebo-Controlled, Phase III Cross-Over Study Evaluating the Oral Neurokinin-1 Antagonist Aprepitant in Combination With a 5HT3 Receptor Antagonist and Dexamethasone in Patients With Germ Cell Tumors Receiving 5-Day Cisplatin Combination Chemotherapy Regimens: A Hoosier Oncology Group Study | Costantine Albany |
| Roisin M. Connolly | Phase I and Pharmacokinetic Study of Romidepsin in Patients with Cancer and Hepatic Dysfunction: A National Cancer Institute Organ DysfunctionWorking Group Study | Combination of Metformin and Gefitinib as First-Line Therapy for Nondiabetic Advanced NSCLC Patients with EGFR Mutations: A Randomized, Double-Blind Phase II Trial | Li Li |
| Priyanka Bhateja | Phase I study of the combination of quinacrine and erlotinib in patients with locally advanced or metastatic non small cell lung cancer | The effect of itraconazole and rifampicin on the pharmacokinetics of osimertinib | Karthick Vishwanathan |
| Jyoti Malhotra | Phase Ib/II study of hydroxychloroquine in combination with chemotherapy in patients with metastatic non-small cell lung cancer (NSCLC) | The effect of itraconazole on the clinical outcomes of patients with advanced non‑small cell lung cancer receiving platinum‑based chemotherapy: a randomized controlled study | Asmaa Waheed Mohamed |
| Oscar Arrieta | Phase II study. Concurrent chemotherapy and radiotherapy with nitroglycerin in locally advanced non-small cell lung cancer | Valproate–doxorubicin: promising therapy for progressing mesothelioma. A phase II study | A. Scherpereel |
| Gregory A. Otterson | Phase II Study of the Histone Deacetylase Inhibitor Romidepsin in Relapsed Small Cell Lung Cancer (Cancer and Leukemia Group B 30304) | Zoledronic acid in patients with stage IIIA/B NSCLC: results of a randomized, phase III study | G. V. Scagliotti |
| Fang Wu | Phase III Randomized Trial of Palonosetron and Dexamethasone With or Without Aprepitant to Prevent Nausea and Vomiting Induced by Fulldose Single-day Cisplatin-based Chemotherapy in Lung Cancer | NCT03861091 | NCT03861091 |
| Xinghao Ai | Pyrotinib combined with thalidomide in advanced non-small-cell lung cancer patients harboring HER2 exon 20 insertions (PRIDE): protocol of an open-label, singlearm phase II trial | NCT00527319 | NCT00527319 |
| Karen L. Reckamp | Randomized Phase 2 Trial of Erlotinib in Combination With High-Dose Celecoxib or Placebo in Patients With Advanced Non-Small Cell Lung Cancer |  |  |
| Tien Hoang | Randomized Phase III Study of Thoracic Radiation in Combination With Paclitaxel and Carboplatin With or Without Thalidomide in Patients With Stage III Non–Small-Cell Lung Cancer: The ECOG 3598 Study |  |  |
| Youngjoo Lee | Randomized Phase II Study of Afatinib Plus Simvastatin Versus Afatinib Alone in Previously Treated Patients with Advanced Nonadenocarcinomatous Non-small Cell Lung Cancer |  |  |
| Oscar Arrieta | Randomized Phase II Trial of All-Trans-Retinoic Acid With Chemotherapy Based on Paclitaxel and Cisplatin As FirstLine Treatment in Patients With Advanced Non–Small-Cell Lung Cancer |  |  |
| Asmaa Waheed Mohamed | The effect of itraconazole on the clinical outcomes of patients with advanced non‑small cell lung cancer receiving platinum‑based chemotherapy: a randomized controlled study |  |  |
| Thierry Berghmans | VAC chemotherapy with valproic acid for refractory/relapsing small cell lung cancer: a phase II study |  |  |
| A. Scherpereel | Valproate–doxorubicin: promising therapy for progressing mesothelioma. A phase II study |  |  |
| G. V. Scagliotti | Zoledronic acid in patients with stage IIIA/B NSCLC: results of a randomized, phase III study |  |  |
| Li Li | Combination of Metformin and Gefitinib as First-Line Therapy for Nondiabetic Advanced NSCLC Patients with EGFR Mutations: A Randomized, Double-Blind Phase II Trial |  |  |
| Xin Shelley Wang | Minocycline Reduces Chemoradiation-Related Symptom Burden in Patients with Non–small Cell Lung Cancer: A Phase II Randomized Trial |  |  |
| Michael J. Seckl | Multicenter, Phase III, Randomized, Double-Blind, PlaceboControlled Trial of Pravastatin Added to First-Line Standard Chemotherapy in Small-Cell Lung Cancer (LUNGSTAR) |  |  |
| Solange Peters | Combined, patient-level, analysis of two randomised trials evaluating the addition of denosumab to standard first-line chemotherapy in advanced NSCLC – The ETOP/EORTC SPLENDOUR and AMGEN-249 trials |  |  |
| Elizabeth A. Johnson | Phase III randomized, double-blind study of maintenance CAI or placebo in patients with advanced non-small cell lung cancer (NSCLC) after completion of initial therapy (NCCTG 97-24-51) |  |  |

**Supplementary Table S9:** List of articles downloaded from Google scholar

| 1 | Goss GD, Jonker DJ, Laurie SA, Weberpals JI, Oza AM, Spaans JN, la Porte C, Dimitroulakos J. A phase I study of high-dose rosuvastatin with standard dose erlotinib in patients with advanced solid malignancies. J Transl Med. 2016 Mar 31;14:83. doi: 10.1186/s12967-016-0836-6. |
| --- | --- |
| 2 | Lacouture ME, Keefe DM, Sonis S, Jatoi A, Gernhardt D, Wang T, Doherty JP, Giri N, Nadanaciva S, O'Connell J, Sbar E, Piperdi B, Garon EB. A phase II study (ARCHER 1042) to evaluate prophylactic treatment of dacomitinib-induced dermatologic and gastrointestinal adverse events in advanced non-small-cell lung cancer. Ann Oncol. 2016 Sep;27(9):1712-8. doi: 10.1093/annonc/mdw227. |
| 3 | Riess JW, Jahchan NS, Das M, Zach Koontz M, Kunz PL, Wakelee HA, Schatzberg A, Sage J, Neal JW. A PHASE IIA STUDY REPOSITIONING DESIPRAMINE IN SMALL CELL LUNG CANCER AND OTHER HIGH-GRADE NEUROENDOCRINE TUMORS. Cancer Treat Res Commun. 2020 Apr 20;23:100174. doi: 10.1016/j.ctarc.2020.100174. |
| 4 | Nechushtan H, Hamamreh Y, Nidal S, Gotfried M, Baron A, Shalev YI, Nisman B, Peretz T, Peylan-Ramu N. A phase IIb trial assessing the addition of disulfiram to chemotherapy for the treatment of metastatic non-small cell lung cancer. Oncologist. 2015 Apr;20(4):366-7. doi: 10.1634/theoncologist.2014-0424. |
| 5 | Marrone KA, Zhou X, Forde PM, Purtell M, Brahmer JR, Hann CL, Kelly RJ, Coleman B, Gabrielson E, Rosner GL, Ettinger DS. A Randomized Phase II Study of Metformin plus Paclitaxel/Carboplatin/Bevacizumab in Patients with Chemotherapy-Naïve Advanced or Metastatic Nonsquamous Non-Small Cell Lung Cancer. Oncologist. 2018 Jul;23(7):859-865. doi: 10.1634/theoncologist.2017-0465. |
| 6 | Dingemans AM, Groen HJ, Herder GJ, Stigt JA, Smit EF, Bahce I, Burgers JA, van den Borne BE, Biesma B, Vincent A, van der Noort V, Aerts JG; NVALT study group. A randomized phase II study comparing paclitaxel-carboplatin-bevacizumab with or without nitroglycerin patches in patients with stage IV nonsquamous nonsmall-cell lung cancer: NVALT12 (NCT01171170)†. Ann Oncol. 2015 Nov;26(11):2286-93. doi: 10.1093/annonc/mdv370. |
| 7 | Seckl MJ, Ottensmeier CH, Cullen M, Schmid P, Ngai Y, Muthukumar D, Thompson J, Harden S, Middleton G, Fife KM, Crosse B, Taylor P, Nash S, Hackshaw A. Multicenter, Phase III, Randomized, Double-Blind, Placebo-Controlled Trial of Pravastatin Added to First-Line Standard Chemotherapy in Small-Cell Lung Cancer (LUNGSTAR). J Clin Oncol. 2017 May 10;35(14):1506-1514. doi: 10.1200/JCO.2016.69.7391. |
| 8 | Keith RL, Blatchford PJ, Merrick DT, Bunn PA Jr, Bagwell B, Dwyer-Nield LD, Jackson MK, Geraci MW, Miller YE. A Randomized Phase II Trial of Pioglitazone for Lung Cancer Chemoprevention in High-Risk Current and Former Smokers. Cancer Prev Res (Phila). 2019 Oct;12(10):721-730. doi: 10.1158/1940-6207.CAPR-19-0006. |
| 9 | Gitlitz BJ, Bernstein E, Santos ES, Otterson GA, Milne G, Syto M, Burrows F, Zaknoen S. A randomized, placebo-controlled, multicenter, biomarker-selected, phase 2 study of apricoxib in combination with erlotinib in patients with advanced non-small-cell lung cancer. J Thorac Oncol. 2014 Apr;9(4):577-82. doi: 10.1097/JTO.0000000000000082. |
| 10 | Musallam KM, Taher AT. Re: Anti-angiogenic therapy using thalidomide combined with chemotherapy in small cell lung cancer: a randomized, double-blind, placebo-controlled trial. J Natl Cancer Inst. 2009 Dec 2;101(23):1657; author reply 1657-8. doi: 10.1093/jnci/djp371. |
| 11 | Hamy AS, Tury S, Wang X, Gao J, Pierga JY, Giacchetti S, Brain E, Pistilli B, Marty M, Espié M, Benchimol G, Laas E, Laé M, Asselain B, Aouchiche B, Edelman M, Reyal F. Celecoxib With Neoadjuvant Chemotherapy for Breast Cancer Might Worsen Outcomes Differentially by COX-2 Expression and ER Status: Exploratory Analysis of the REMAGUS02 Trial. J Clin Oncol. 2019 Mar 10;37(8):624-635. doi: 10.1200/JCO.18.00636. |
| 12 | Rengan R, Mick R, Pryma DA, Lin LL, Christodouleas J, Plastaras JP, Simone CB 2nd, Gupta AK, Evans TL, Stevenson JP, Langer CJ, Kucharczuk J, Friedberg J, Lam S, Patsch D, Hahn SM, Maity A. Clinical Outcomes of the HIV Protease Inhibitor Nelfinavir With Concurrent Chemoradiotherapy for Unresectable Stage IIIA/IIIB Non-Small Cell Lung Cancer: A Phase 1/2 Trial. JAMA Oncol. 2019 Oct 1;5(10):1464-1472. doi: 10.1001/jamaoncol.2019.2095. |
| 13 | Dilly SJ, Morris GS, Taylor PC, Parmentier F, Williams C, Afshar M. Clinical Pharmacokinetics of a Lipid-Based Formulation of Risperidone, VAL401: Analysis of a Single Dose in an Open-Label Trial of Late-Stage Cancer Patients. Eur J Drug Metab Pharmacokinet. 2019 Aug;44(4):557-565. doi: 10.1007/s13318-018-00538-4. |
| 14 | Gerber DE, Putnam WC, Fattah FJ, Kernstine KH, Brekken RA, Pedrosa I, Skelton R, Saltarski JM, Lenkinski RE, Leff RD, Ahn C, Padmanabhan C, Chembukar V, Kasiri S, Kallem RR, Subramaniyan I, Yuan Q, Do QN, Xi Y, Reznik SI, Pelosof L, Faubert B, DeBerardinis RJ, Kim J. Concentration-dependent Early Antivascular and Antitumor Effects of Itraconazole in Non-Small Cell Lung Cancer. Clin Cancer Res. 2020 Nov 15;26(22):6017-6027. doi: 10.1158/1078-0432.CCR-20-1916. |
| 15 | Hu MI, Glezerman IG, Leboulleux S, Insogna K, Gucalp R, Misiorowski W, Yu B, Zorsky P, Tosi D, Bessudo A, Jaccard A, Tonini G, Ying W, Braun A, Jain RK. Denosumab for treatment of hypercalcemia of malignancy. J Clin Endocrinol Metab. 2014 Sep;99(9):3144-52. doi: 10.1210/jc.2014-1001. |
| 16 | Deplanque G, Gervais R, Vergnenegre A, Falchero L, Souquet PJ, Chavaillon JM, Taviot B, Fraboulet G, Saal H, Robert C, Chosidow O; CYTAR investigators. Doxycycline for prevention of erlotinib-induced rash in patients with non-small-cell lung cancer (NSCLC) after failure of first-line chemotherapy: A randomized, open-label trial. J Am Acad Dermatol. 2016 Jun;74(6):1077-85. doi: 10.1016/j.jaad.2016.01.019. |
| 17 | Koch A, Bergman B, Holmberg E, Sederholm C, Ek L, Kosieradzki J, Lamberg K, Thaning L, Ydreborg SO, Sörenson S; Swedish Lung Cancer Study Group. Effect of celecoxib on survival in patients with advanced non-small cell lung cancer: a double blind randomised clinical phase III trial (CYCLUS study) by the Swedish Lung Cancer Study Group. Eur J Cancer. 2011 Jul;47(10):1546-55. doi: 10.1016/j.ejca.2011.03.035. |
| 18 | Edelman MJ, Wang X, Hodgson L, Cheney RT, Baggstrom MQ, Thomas SP, Gajra A, Bertino E, Reckamp KL, Molina J, Schiller JH, Mitchell-Richards K, Friedman PN, Ritter J, Milne G, Hahn OM, Stinchcombe TE, Vokes EE; Alliance for Clinical Trials in Oncology. Phase III Randomized, Placebo-Controlled, Double-Blind Trial of Celecoxib in Addition to Standard Chemotherapy for Advanced Non-Small-Cell Lung Cancer With Cyclooxygenase-2 Overexpression: CALGB 30801 (Alliance). J Clin Oncol. 2017 Jul 1;35(19):2184-2192. doi: 10.1200/JCO.2016.71.3743. |
| 19 | Bi N, Liang J, Zhou Z, Chen D, Fu Z, Yang X, Feng Q, Hui Z, Xiao Z, Lv J, Wang X, Zhang T, Wang X, Deng L, Wang W, Wang J, Liu L, Hu C, Wang L. Effect of Concurrent Chemoradiation With Celecoxib vs Concurrent Chemoradiation Alone on Survival Among Patients With Non-Small Cell Lung Cancer With and Without Cyclooxygenase 2 Genetic Variants: A Phase 2 Randomized Clinical Trial. JAMA Netw Open. 2019 Dec 2;2(12):e1918070. doi: 10.1001/jamanetworkopen.2019.18070. |
| 20 | Garland LL, Guillen-Rodriguez J, Hsu CH, Yozwiak M, Zhang HH, Alberts DS, Davis LE, Szabo E, Merenstein C, Lel J, Zhang X, Liu H, Liu G, Spira AE, Beane JE, Wojtowicz M, Chow HS. Effect of Intermittent Versus Continuous Low-Dose Aspirin on Nasal Epithelium Gene Expression in Current Smokers: A Randomized, Double-Blinded Trial. Cancer Prev Res (Phila). 2019 Nov;12(11):809-820. doi: 10.1158/1940-6207.CAPR-19-0036. |
| 21 | Gridelli C, Gallo C, Ceribelli A, Gebbia V, Gamucci T, Ciardiello F, Carozza F, Favaretto A, Daniele B, Galetta D, Barbera S, Rosetti F, Rossi A, Maione P, Cognetti F, Testa A, Di Maio M, Morabito A, Perrone F; GECO investigators. Factorial phase III randomised trial of rofecoxib and prolonged constant infusion of gemcitabine in advanced non-small-cell lung cancer: the GEmcitabine-COxib in NSCLC (GECO) study. Lancet Oncol. 2007 Jun;8(6):500-12. doi: 10.1016/S1470-2045(07)70146-8. |
| 22 | Vishwanathan K, Cantarini M, So K, Masson E, Fetterolf J, Ramalingam SS, Harvey RD. Impact of Disease and Treatment Response in Drug-Drug Interaction Studies: Osimertinib and Simvastatin in Advanced Non-Small Cell Lung Cancer. Clin Transl Sci. 2020 Jan;13(1):41-46. doi: 10.1111/cts.12688. |
| 23 | Bonanni B, Serrano D, Maisonneuve P, Veronesi G, Johansson H, Aristarco V, Varricchio C, Cazzaniga M, Lazzeroni M, Rampinelli C, Bellomi M, Vecchi M, Spaggiari L, Vornik L, Brown PH, Beavers T, Guerrieri-Gonzaga A, Szabo E. Low-Dose Aspirin in High-Risk Individuals With Screen-Detected Subsolid Lung Nodules: A Randomized Phase II Trial. JNCI Cancer Spectr. 2020 Oct 20;4(6):pkaa096. doi: 10.1093/jncics/pkaa096. |
| 24 | Mao JT, Roth MD, Fishbein MC, Aberle DR, Zhang ZF, Rao JY, Tashkin DP, Goodglick L, Holmes EC, Cameron RB, Dubinett SM, Elashoff R, Szabo E, Elashoff D. Lung cancer chemoprevention with celecoxib in former smokers. Cancer Prev Res (Phila). 2011 Jul;4(7):984-93. doi: 10.1158/1940-6207.CAPR-11-0078. |
| 25 | Skwarski M, McGowan DR, Belcher E, Di Chiara F, Stavroulias D, McCole M, Derham JL, Chu KY, Teoh E, Chauhan J, O'Reilly D, Harris BHL, Macklin PS, Bull JA, Green M, Rodriguez-Berriguete G, Prevo R, Folkes LK, Campo L, Ferencz P, Croal PL, Flight H, Qi C, Holmes J, O'Connor JPB, Gleeson FV, McKenna WG, Harris AL, Bulte D, Buffa FM, Macpherson RE, Higgins GS. Mitochondrial Inhibitor Atovaquone Increases Tumor Oxygenation and Inhibits Hypoxic Gene Expression in Patients with Non-Small Cell Lung Cancer. Clin Cancer Res. 2021 May 1;27(9):2459-2469. doi: 10.1158/1078-0432.CCR-20-4128. |
| 26 | Arrieta O, Hernández-Pedro N, Maldonado F, Ramos-Ramírez M, Yamamoto-Ramos M, López-Macías D, Lozano F, Zatarain-Barrón ZL, Turcott JG, Barrios-Bernal P, Orozco-Morales M, Flores-Estrada D, Cardona AF, Rolfo C, Cacho-Díaz B. Nitroglycerin Plus Whole Intracranial Radiation Therapy for Brain Metastases in Patients With Non-Small Cell Lung Cancer: A Randomized, Open-Label, Phase 2 Clinical Trial. Int J Radiat Oncol Biol Phys. 2023 Mar 1;115(3):592-607. doi: 10.1016/j.ijrobp.2022.02.010. |
| 27 | T. Tsakiridis, P. Ellis, J.C. Cutz, G. Pond, M. Wierzbicki, P. Kavsak, J. Wright, OCOG-ALMERA: A phase II trial investigating the ability of metformin to chemo-radio-sensitize and prevent recurrence in locally advanced (LA) non-small cell lung cancer (NSCLC). J. Thorac. Oncol., 11 (2016), pp. S50-S51 |
| 28 | Owonikoko TK, Zhang G, Kim HS, Stinson RM, Bechara R, Zhang C, Chen Z, Saba NF, Pakkala S, Pillai R, Deng X, Sun SY, Rossi MR, Sica GL, Ramalingam SS, Khuri FR. Patient-derived xenografts faithfully replicated clinical outcome in a phase II co-clinical trial of arsenic trioxide in relapsed small cell lung cancer. J Transl Med. 2016 May 3;14(1):111. doi: 10.1186/s12967-016-0861-5. |
| 29 | Su JM, Li XN, Thompson P, Ou CN, Ingle AM, Russell H, Lau CC, Adamson PC, Blaney SM. Phase 1 study of valproic acid in pediatric patients with refractory solid or CNS tumors: a children's oncology group report. Clin Cancer Res. 2011 Feb 1;17(3):589-97. doi: 10.1158/1078-0432.CCR-10-0738. |
| 30 | Rudin CM, Brahmer JR, Juergens RA, Hann CL, Ettinger DS, Sebree R, Smith R, Aftab BT, Huang P, Liu JO. Phase 2 study of pemetrexed and itraconazole as second-line therapy for metastatic nonsquamous non-small-cell lung cancer. J Thorac Oncol. 2013 May;8(5):619-23. doi: 10.1097/JTO.0b013e31828c3950. |
| 31 | Connolly RM, Laille E, Vaishampayan U, Chung V, Kelly K, Dowlati A, Alese OB, Harvey RD, Haluska P, Siu LL, Kummar S, Piekarz R, Ivy SP, Anders NM, Downs M, O'Connor A, Scardina A, Saunders J, Rosner GL, Carducci MA, Rudek MA; ETCTN-9008 Study Team. Phase I and Pharmacokinetic Study of Romidepsin in Patients with Cancer and Hepatic Dysfunction: A National Cancer Institute Organ Dysfunction Working Group Study. Clin Cancer Res. 2020 Oct 15;26(20):5329-5337. doi: 10.1158/1078-0432.CCR-20-1412. |
| 32 | Bhateja P, Dowlati A, Sharma N. Phase I study of the combination of quinacrine and erlotinib in patients with locally advanced or metastatic non small cell lung cancer. Invest New Drugs. 2018 Jun;36(3):435-441. doi: 10.1007/s10637-017-0515-3. |
| 33 | Malhotra J, Jabbour S, Orlick M, Riedlinger G, Guo Y, White E, Aisner J. Phase Ib/II study of hydroxychloroquine in combination with chemotherapy in patients with metastatic non-small cell lung cancer (NSCLC). Cancer Treat Res Commun. 2019;21:100158. doi: 10.1016/j.ctarc.2019.100158. |
| 34 | Arrieta O, Blake M, de la Mata-Moya MD, Corona F, Turcott J, Orta D, Alexander-Alatorre J, Gallardo-Rincón D. Phase II study. Concurrent chemotherapy and radiotherapy with nitroglycerin in locally advanced non-small cell lung cancer. Radiother Oncol. 2014 May;111(2):311-5. doi: 10.1016/j.radonc.2014.01.021. |
| 35 | Otterson GA, Hodgson L, Pang H, Vokes EE; Cancer and Leukemia Group B. Phase II study of the histone deacetylase inhibitor Romidepsin in relapsed small cell lung cancer (Cancer and Leukemia Group B 30304). J Thorac Oncol. 2010 Oct;5(10):1644-8. doi: 10.1097/JTO.0b013e3181ec1713. |
| 36 | Wu F, Lin X, Yang Z, Sun Z, Zeng F, Heng J, Qu J, Zeng L, Yang N, Zhang Y. Phase III Randomized Trial of Palonosetron and Dexamethasone With or Without Aprepitant to Prevent Nausea and Vomiting Induced by Full-dose Single-day Cisplatin-based Chemotherapy in Lung Cancer. Clin Lung Cancer. 2018 Nov;19(6):e913-e918. doi: 10.1016/j.cllc.2018.08.006. |
| 37 | Ai X, Song Z, Jian H, Zhou Z, Chen Z, Yu Y, Li Z, Lu S. Pyrotinib combined with thalidomide in advanced non-small-cell lung cancer patients harboring HER2 exon 20 insertions (PRIDE): protocol of an open-label, single-arm phase II trial. BMC Cancer. 2021 Sep 16;21(1):1033. doi: 10.1186/s12885-021-08759-8. |
| 38 | Reckamp KL, Koczywas M, Cristea MC, Dowell JE, Wang HJ, Gardner BK, Milne GL, Figlin RA, Fishbein MC, Elashoff RM, Dubinett SM. Randomized phase 2 trial of erlotinib in combination with high-dose celecoxib or placebo in patients with advanced non-small cell lung cancer. Cancer. 2015 Sep 15;121(18):3298-306. doi: 10.1002/cncr.29480. |
| 39 | Hoang T, Dahlberg SE, Schiller JH, Mehta MP, Fitzgerald TJ, Belinsky SA, Johnson DH. Randomized phase III study of thoracic radiation in combination with paclitaxel and carboplatin with or without thalidomide in patients with stage III non-small-cell lung cancer: the ECOG 3598 study. J Clin Oncol. 2012 Feb 20;30(6):616-22. doi: 10.1200/JCO.2011.36.9116. |
| 40 | Lee Y, Lee KH, Lee GK, Lee SH, Lim KY, Joo J, Go YJ, Lee JS, Han JY. Randomized Phase II Study of Afatinib Plus Simvastatin Versus Afatinib Alone in Previously Treated Patients with Advanced Nonadenocarcinomatous Non-small Cell Lung Cancer. Cancer Res Treat. 2017 Oct;49(4):1001-1011. doi: 10.4143/crt.2016.546. |
| 41 | Arrieta O, González-De la Rosa CH, Aréchaga-Ocampo E, Villanueva-Rodríguez G, Cerón-Lizárraga TL, Martínez-Barrera L, Vázquez-Manríquez ME, Ríos-Trejo MA, Alvarez-Avitia MA, Hernández-Pedro N, Rojas-Marín C, De la Garza J. Randomized phase II trial of All-trans-retinoic acid with chemotherapy based on paclitaxel and cisplatin as first-line treatment in patients with advanced non-small-cell lung cancer. J Clin Oncol. 2010 Jul 20;28(21):3463-71. doi: 10.1200/JCO.2009.26.6452. |
| 42 | Mohamed AW, Elbassiouny M, Elkhodary DA, Shawki MA, Saad AS. The effect of itraconazole on the clinical outcomes of patients with advanced non-small cell lung cancer receiving platinum-based chemotherapy: a randomized controlled study. Med Oncol. 2021 Feb 9;38(3):23. doi: 10.1007/s12032-021-01475-0. |
| 43 | Berghmans T, Lafitte JJ, Scherpereel A, Ameye L, Paesmans M, Meert AP, Colinet B, Tulippe C, Willems L, Leclercq N, Sculier JP; European Lung Cancer Working Party. VAC chemotherapy with valproic acid for refractory/relapsing small cell lung cancer: a phase II study. ERJ Open Res. 2015 Oct 19;1(2):00029-2015. doi: 10.1183/23120541.00029-2015. |
| 44 | Scherpereel A, Berghmans T, Lafitte JJ, Colinet B, Richez M, Bonduelle Y, Meert AP, Dhalluin X, Leclercq N, Paesmans M, Willems L, Sculier JP; European Lung Cancer Working Party (ELCWP). Valproate-doxorubicin: promising therapy for progressing mesothelioma. A phase II study. Eur Respir J. 2011 Jan;37(1):129-35. doi: 10.1183/09031936.00037310. |
| 45 | Scagliotti GV, Kosmidis P, de Marinis F, Schreurs AJM, Albert I, Engel-Riedel W, Schallier D, Barbera S, Kuo HP, Sallo V, Perez JR, Manegold C. Zoledronic acid in patients with stage IIIA/B NSCLC: results of a randomized, phase III study. Ann Oncol. 2012 Aug;23(8):2082-2087. doi: 10.1093/annonc/mds128. |
| 46 | Daniel Shao, Weng Tan, 532TiP - CANOPY-1: A phase III, placebo-controlled study of pembrolizumab (PEM) plus platinum-based doublet chemotherapy (Ctx) with/without canakinumab in untreated patients (pts) with stage IIIB/IIIC-IV NSCLC. Annals of Oncology (2019) 30 (suppl_9): ix157-ix181. 10.1093/annonc/mdz437 |
| 47 | D. Lim, Y. Goto, B.C. Cho, H. Kaneda, J.-H. Kang, S.-W. Kim, C.- H. Yang, W.C. Su, K. Obyrne, H. Chiu, J.C. V. Papadimitrakopoulou, M. Reck, I. Malet, B. Mookerjee, Z. Zewen, L. Paz-Ares Rodriguez, 533TiP - CANOPY-2 A phase III, placebo-controlled study of canakinumab with or without docetaxel in patients (pts) with NSCLC previously treated with PD-(L)1 inhibitors and platinum-based chemotherapy (Ctx). Annals of Oncology (2019) 30, (Supplement 9): ix179-ix180 doi.org/10.1093/annonc/mdz437.058 |
| 48 | A. Santoro, P. Garrido Lopez, D.S.W. Tan, L. Paz-Ares, F. Shepherd, A. Bearz, F. Barlesi, J.F. Vansteenkiste, T.M. Kim, T.R. Overbeck, I.I. Rybkin, E. Felip, W. Zhou, L. Santarpia, S. Eddy, E.S. Schaefer, 1575P - Preliminary results from phase Ib study of spartalizumab plus chemotherapy for advanced non-small cell lung cancer (NSCLC). Annals of Oncology (2019) 30, (Supplement 5) doi:10.1093/annonc/mdz260 |
| 49 | Tilves C, Yeh HC, Maruthur N, Juraschek SP, Miller ER, Appel LJ, Mueller NT. A behavioral weight-loss intervention, but not metformin, decreases a marker of gut barrier permeability: results from the SPIRIT randomized trial. Int J Obes (Lond). 2022 Mar;46(3):655-660. doi: 10.1038/s41366-021-01039-2. |
| 50 | Li KL, Li L, Zhang P, Kang J, Wang YB, Chen HY, He Y. A Multicenter Double-blind Phase II Study of Metformin With Gefitinib as First-line Therapy of Locally Advanced Non-Small-cell Lung Cancer. Clin Lung Cancer. 2017 May;18(3):340-343. doi: 10.1016/j.cllc.2016.12.003. |
| 51 | Cho BC, Goldberg SB, Kim DW, Socinski MA, Burns TF, Lwin Z, Pathan N, Ma WD, Masters JC, Cossons N, Wilner K, Nishio M, Husain H. A phase 1b/2 study of PF-06747775 as monotherapy or in combination with Palbociclib in patients with epidermal growth factor receptor mutant advanced non-small cell lung cancer. Expert Opin Investig Drugs. 2022 Jul;31(7):747-757. doi: 10.1080/13543784.2022.2075341. |
| 52 | [Andrea Borghese Apolo](https://ascopubs.org/author/Apolo%2C+Andrea+Borghese) , [Howard L. Parnes](https://ascopubs.org/author/Parnes%2C+Howard+L) , [Ravi Amrit Madan](https://ascopubs.org/author/Madan%2C+Ravi+Amrit) , [James L. Gulley](https://ascopubs.org/author/Gulley%2C+James+L) , [Jane B. Trepel](https://ascopubs.org/author/Trepel%2C+Jane+B) , [Min-Jung Lee](https://ascopubs.org/author/Lee%2C+Min-Jung)[Sunmin Lee](https://ascopubs.org/author/Lee%2C+Sunmin) , [Seth M. Steinberg](https://ascopubs.org/author/Steinberg%2C+Seth+M) , [Simone John](https://ascopubs.org/author/JOHN%2C+Simone) , [Sylvia Vania Alarcon Velasco](https://ascopubs.org/author/Alarcon+Velasco%2C+Sylvia+Vania) , [William Douglas Figg](https://ascopubs.org/author/Figg%2C+William+Douglas) , [William L. Dahut](https://ascopubs.org/author/Dahut%2C+William+L), A phase I study of gemcitabine, carboplatin, and lenalidomide for treatment of patients with advanced metastatic urothelial carcinoma (UC) and other solid tumors. Journal of Clinical Oncology (2014) 32, no. 15_suppl. DOI: 10.1200/jco.2014.32.15_suppl.e15527 |
| 53 | Jamil MO, Jerome MS, Miley D, Selander KS, Robert F. A pilot study of zoledronic acid in the treatment of patients with advanced malignant pleural mesothelioma. Lung Cancer (Auckl). 2017 Jun 12;8:39-44. doi: 10.2147/LCTT.S135802. |
| 54 | D.R. Spigel, F.R. Hirsch, R.H. De Boer, R. Natale, J. Crawford, G.J. Weiss, J. Glaspy, A. Feng, A. Braun, R. Jain, A randomized, double-blind, multicenter phase 2 trial of denosumab in combination with chemotherapy as first-line treatment of metastatic non-small cell lung cancer. Annals of Oncology (2014) 25 (Supple4) DOI:https://doi.org/10.1093/annonc/mdu349.109 |
| 55 | Claudio Vernieri; Federico Nichetti; Francesca Ligorio; Emma Zattarin; Teresa Beninato; Riccardo Lobefaro; Giulia Bianchi; Giuseppe Capri; Marina Garassino; Giuseppe Lo Russo; Michele Del Vecchio; Paola Corsetto; Licia Rivoltini; Chiara Castelli; Filippo de Braud, Abstract CT198 Efficacy of metformin in Preventing glucocorticoid-induced diabetes in Melanoma, breast or Lung Cancer patients with brain metastases The phase II OPTIMAL study. Cancer Res (2020) 80 (16_Supplement): CT198. [doi.org/10.1158/1538-7445.AM2020-CT198](https://doi.org/10.1158/1538-7445.AM2020-CT198) |
| 56 | Luis Paz-Ares; Edward B. Garon; Tony Mok; Andrea Ardizzoni; Fabrice Barlesi; Byoung Chul Cho; Gilberto de Castro; Pedro De Marchi; Enriqueta Felip; Yasushi Goto; Alastair Greystoke; Shun Lu; Darren Wan-Teck Lim; Martin Reck; Benjamin J. Solomon; David R. Spigel; Daniel SW Tan; Michael Thomas; James Chih-Hsin Yang; Jay M. Lee; Pilar Garrido; Edward Kim; Bruce E. Johnson, CANOPY program clinical trials Canakinumab (Cana) in patients (pts) with non-small cell lung cancer (NSCLC). Cancer Res (2020) 80 (16_Supplement): CT286. [doi.org/10.1158/1538-7445.AM2020-CT286](https://doi.org/10.1158/1538-7445.AM2020-CT286) |
| 57 | Lotte Engell-Noerregaard, Pia Kvistborg, Mai-Britt Zocca, Ayako W. Pedersen, Mogens H. Claesson, Anders Mellemgaard, Clinical and Immunological Effects in Patients with Advanced Non-Small Cell Lung-Cancer after Vaccination with Dendritic Cells Exposed to an Allogeneic Tumor Cell Lysate. [World Journal of Vaccines](https://www.scirp.org/journal/journalarticles.aspx?journalid=510) (2013) [3 (2),](https://www.scirp.org/journal/home.aspx?issueid=2984#31520) DOI: [10.4236/wjv.2013.32011](http://dx.doi.org/10.4236/wjv.2013.32011) |
| 58 | Oscar Gerardo Arrieta Rodriguez, Feliciano Barron Barron, Miguel-Ángel Salinas Padilla, Laura Alejandra Ramirez-Tirado, Diana Flores-Estrada, Graciela Cruz-Rico, Manuel Jesús Arguelles Jiménez, Andres Felipe Cardona Zorrilla, Combination of metformin plus TKI vs. TKI alone in EGFR(+) LUNG adenocarcinoma A randomized phase II study. (2018) Journal of Clinical Oncology 36, no. 15_suppl 9013-9013. DOI: 10.1200/JCO.2018.36.15_suppl.9013 |
| 59 | Helen Ross, Verline Justilien, Alan Fields, Combined PKCi and mTOR Inhibition in Advanced or Recurrent Lung Cancer Preliminary Report of an Ongoing Phase I II Trial. Journal of Thoracic Oncology (2017), 12 (1), S1074. doi.org/10.1016/j.jtho.2016.11.1502 |
| 60 | Zhou T, Zhang Y, Ma Y, Ma W, Wu X, Huang L, Feng W, Zhou H, Liu J, Zhao H, Zhang L, Yang Y, Huang Y. Comparison of aprepitant versus desloratadine for EGFR-TKI-induced pruritus: A randomized phase 2 clinical trial. Cancer. 2022 Nov 15;128(22):3969-3976. doi: 10.1002/cncr.34474. |
| 61 | D. Ross Camidge, Gregory Alan Otterson, Jeffrey W. Clark, Sai-Hong Ignatius Ou, Jared Weiss, Steven Ades, Umberto Conte, Yiyun Tang, Sherry Chia-E Wang, Danielle Murphy, Keith D. Wilner, Liza Cosca Villaruz, Crizotinib in patients (pts) with MET-amplified non-small cell lung cancer (NSCLC) Updated safety and efficacy findings from a phase 1 trial. Journal of Clinical Oncology (2018) 36, (suppl 15) 9062-9062. DOI: 10.1200/JCO.2018.36.15 |
| 62 | Maeng CH, Kim BH, Chon J, Kang WS, Kang K, Woo M, Hong IK, Lee J, Lee KY. Effect of multimodal intervention care on cachexia in patients with advanced cancer compared to conventional management (MIRACLE): an open-label, parallel, randomized, phase 2 trial. Trials. 2022 Apr 11;23(1):281. doi: 10.1186/s13063-022-06221-z. |
| 63 | Harvey RD, Aransay NR, Isambert N, Lee JS, Arkenau T, Vansteenkiste J, Dickinson PA, Bui K, Weilert D, So K, Thomas K, Vishwanathan K. Effect of multiple-dose osimertinib on the pharmacokinetics of simvastatin and rosuvastatin. Br J Clin Pharmacol. 2018 Dec;84(12):2877-2888. doi: 10.1111/bcp.13753. |
| 64 | Y. Lee, S.-H. Lee, G.K. Lee, E.J. Lim, J.-Y. Han, EP14.01-023 A Randomized Phase II Study of Irinotecan Plus Cisplatin With or Without Simvastatin in Ever-Smoking Small Cell Lung Cancer. Journal of Thoracic Oncology (2022), 17, (Supplement 9), S535-S536. <https://doi.org/10.1016/j.jtho.2022.07.958> |
| 65 | Scagliotti, G., Manegold, C., de Marinis, F., Engel-Riedel, W., Albert, I., Sallo, V., Chen, Y.M., Perez, J.R., Kosmidis, P. Evaluating the efficacy of zoledronic acid for the prevention of disease progression in patients with non-small cell lung cancer (NSCLC). European Journal of Cancer Supplements (2009) 7(2):522-522 DOI: 10.1016/S1359-6349(09)71769-0 |
| 66 | Marco Giallombardo, Jorge Jorge Chacartegui, Pablo Reclusa, Jan P. Van Meerbeeck, Riccardo Alessandro, Marc Peeters, Patrick Pauwels, Christian Diego Rolfo, follow up analysis by exosomal miRNAs in EGFR mutated non-small cell lung cancer (NSCLC) patients during osimertinib (AZD9291) treatment A potential prognostic biomarker tool. Journal of Clinical Oncology (2016) 34, 15_suppl. DOI: 10.1200/JCO.2016.34.15_suppl.e23035 |
| 67 | Pichon B, Campion L, Delpon G, Thillays F, Carrie C, Cellier P, Pommier P, Laude C, Mervoyer A, Hamidou H, Mahé MA, Supiot S. High-Dose Hypofractionated Radiation Therapy for Noncompressive Vertebral Metastases in Combination With Zoledronate: A Phase 1 Study. Int J Radiat Oncol Biol Phys. 2016 Nov 15;96(4):840-847. doi: 10.1016/j.ijrobp.2016.07.027. |
| 68 | Wink KC, Belderbos JS, Dieleman EM, Rossi M, Rasch CR, Damhuis RA, Houben RM, Troost EG. Improved progression free survival for patients with diabetes and locally advanced non-small cell lung cancer (NSCLC) using metformin during concurrent chemoradiotherapy. Radiother Oncol. 2016 Mar;118(3):453-9. doi: 10.1016/j.radonc.2016.01.012. |
| 69 | Skinner H, Hu C, Tsakiridis T, Santana-Davila R, Lu B, Erasmus JJ, Doemer AJ, Videtic GMM, Coster J, Yang AX, Lee RY, Werner-Wasik M, Schaner PE, McCormack SE, Esparaz BT, McGarry RC, Bazan J, Struve T, Paulus R, Bradley JD. Addition of Metformin to Concurrent Chemoradiation in Patients With Locally Advanced Non-Small Cell Lung Cancer: The NRG-LU001 Phase 2 Randomized Clinical Trial. JAMA Oncol. 2021 Sep 1;7(9):1324-1332. doi: 10.1001/jamaoncol.2021.2318. |
| 70 | T.S.K. Mok, J-L. Pujol, M. Tsuboi, J. Lee, E. Kim, O. Leonov, J. Zhang, J. Duan, C. Lobetti-Bodoni, J.C. Brase, A. Savchenko, P. Garrido Lopez, LBA4 CANOPY-N A phase II study of canakinumab (CAN) or pembrolizumab (PEM), alone or in combination, as neoadjuvant therapy in patients (pts) with resectable stage Ib–IIIa non-small cell lung cancer (NSCLC). Annals of Oncology, (2022) 33, (SUPPLEMENT 9), S1547-S1548 DOI:https://doi.org/10.1016/j.annonc.2022.10.322 |
| 71 | E.B. Garon, S. Lu, Y. Goto, P.R. De Marchi, L. Paz-Ares, D.R. Spigel, M. Thomas, J.C-H. Yang, A. Ardizzoni, F. Barlesi, S. Khanna, C. Bossen, M. Carbini, A. Yovine, B.C. Cho, LBA49 CANOPY-A Phase III study of canakinumab (CAN) as adjuvant therapy in patients (pts) with completely resected non-small cell lung cancer (NSCLC). Annals of Oncology, (2022) 33, SUPPLEMENT 7, S1414-S1415 DOI:https://doi.org/10.1016/j.annonc.2022.08.049 |
| 72 | Rui Han, Jianghua Li, Yubo Wang, Tingting He, Jie Zheng, Yong He, Low BMI patients with advanced EGFR mutation-positive NSCLC can get a better outcome from metformin plus EGFR-TKI as first-line therapy A secondary analysis of a phase 2 randomized clinical trial. Chinese Medical Journal Pulmonary and Critical Care Medicine, (2023) 1, (2), Pages 119-124. <https://doi.org/10.1016/j.pccm.2023.04.006> |
| 73 | Chun SG, Liao Z, Jeter MD, Chang JY, Lin SH, Komaki RU, Guerrero TM, Mayo RC, Korah BM, Koshy SM, Heymach JV, Koong AC, Skinner HD. Metabolic Responses to Metformin in Inoperable Early-stage Non-Small Cell Lung Cancer Treated With Stereotactic Radiotherapy: Results of a Randomized Phase II Clinical Trial. Am J Clin Oncol. 2020 Apr;43(4):231-235. doi: 10.1097/COC.0000000000000632. |
| 74 | Wang XS, Shi Q, Mendoza T, Lin S, Chang JY, Bokhari RH, Lin HK, Garcia-Gonzalez A, Kamal M, Cleeland CS, Liao Z. Minocycline Reduces Chemoradiation-Related Symptom Burden in Patients with Non-Small Cell Lung Cancer: A Phase 2 Randomized Trial. Int J Radiat Oncol Biol Phys. 2020 Jan 1;106(1):100-107. doi: 10.1016/j.ijrobp.2019.10.010. |
| 75 | M. Di Maio, M.C. Piccirillo, G. Daniele, F. Nuzzo, C. Gridelli, V. Gebbia, F. Ciardiello, S. De Placido, A. Ceribelli, A. Favaretto, A. De Matteis, R. Feld, C.A. Butts, N.B. Leighl, A. Morabito, J. Bryce, S. Signoriello, C. Gallo, F. Perrone, Symptomatic Toxicities Experienced During Anti-cancer Treatment: Comparison of Patients’ and Physicians’ Reporting in Three Randomized Controlled Trials (RSCT). 2014, 25, SUPPLEMENT 4, IV517. DOI:https://doi.org/10.1093/annonc/mdu356.2 |
| 76 | Reymen BJT, van Gisbergen MW, Even AJG, Zegers CML, Das M, Vegt E, Wildberger JE, Mottaghy FM, Yaromina A, Dubois LJ, van Elmpt W, De Ruysscher D, Lambin P. Nitroglycerin as a radiosensitizer in non-small cell lung cancer: Results of a prospective imaging-based phase II trial. Clin Transl Radiat Oncol. 2019 Dec 13;21:49-55. doi: 10.1016/j.ctro.2019.12.002. |
| 77 | Murtaza M, Dawson SJ, Tsui DW, Gale D, Forshew T, Piskorz AM, Parkinson C, Chin SF, Kingsbury Z, Wong AS, Marass F, Humphray S, Hadfield J, Bentley D, Chin TM, Brenton JD, Caldas C, Rosenfeld N. Non-invasive analysis of acquired resistance to cancer therapy by sequencing of plasma DNA. Nature. 2013 May 2;497(7447):108-12. doi: 10.1038/nature12065. |
| 78 | Catherine H. Van Poznak, Joseph M. Unger, Amy K. Darke, Carol Moinpour, Robert A. Bagramian, Mark M. Schubert, Lisa Kathryn Hansen, Justin D. Floyd, Shaker R. Dakhil, Danika L. Lew, James Lloyd Wade, Michael Jordan Fisch, Norah Lynn Henry, Dawn L. Hershman, Julie Gralow, Osteonecrosis of the jaw in patients with cancer receiving zoledronic acid for bone metastases SWOG S0702, NCT00874211. Journal of Clinical Oncology 37, no. 15_suppl (May 20, 2019) 11502-11502. DOI: 10.1200/JCO.2019.37.15_suppl.11502 |
| 79 | O. Arrieta, J. Turcott, F. Barrón, M. Ramos, S. Yendamuri, L. Zatarain Barron, A. Cardona Zorrilla, R. Rosell , P48.09 Body Mass Index Predicts Benefit From Adding Metformin to EGFR-TKIs in Patients With Lung Adenocarcinoma Subanalysis From an Randomized controlled trial. Journal of Thoracic Oncology (2021) 16, (10 SUPPLEMENT), S1109-S1110 DOI:https://doi.org/10.1016/j.jtho.2021.08.520 |
| 80 | L. Wu, K. Li, B. Chen, W. Peng, J. Wang, M. Jiang, Q. Wang, X. Pu, J. Li, F. Xu, Y. Xu, P48.15 A Case from a Single-Arm, Phase Two, Open Label Study Assessing Sindilimab Plus Metaformin in Chemotherapy Failed PD-L1 Positive Advanced SCLC. Journal of Thoracic Oncology 2021, 16, (SUPPLEMENT 3), S505-S506 DOI:https://doi.org/10.1016/j.jtho.2021.01.885 |
| 81 | Li L, Jiang L, Wang Y, Zhao Y, Zhang XJ, Wu G, Zhou X, Sun J, Bai J, Ren B, Tian K, Xu Z, Xiao HL, Zhou Q, Han R, Chen H, Wang H, Yang Z, Gao C, Cai S, He Y. Combination of Metformin and Gefitinib as First-Line Therapy for Nondiabetic Advanced NSCLC Patients with EGFR Mutations: A Randomized, Double-Blind Phase II Trial. Clin Cancer Res. 2019 Dec 1;25(23):6967-6975. doi: 10.1158/1078-0432.CCR-19-0437. |
| 82 | David E. Gerber, Rachael Skelton, Ying Dong, Laurin Loudat, Jonathan Dowell, David A. BoothmanVenetia Sarode, Wei Zhang, Yang Xie, Adi Gazdar, Eugene P. Frenkel, Joan H. Schiller, Phase I and pharmacodynamic study of the histone deacetylase (HDAC) inhibitor romidepsin plus erlotinib in previously treated advanced non-small cell lung cancer (NSCLC). Journal of Clinical Oncology 31, no. 15_suppl (May 20, 2013) 8088-8088. DOI: 10.1200/jco.2013.31.15_suppl.8088 |
| 83 | Dennis A. Wigle , Sumithra J. Mandrekar , Katie Allen-Ziegler , Yaron Gesthalter , Paul Holland , Marie-Christine AubryPaul J. Limburg , Spira Avi , Eva Szabo, Pioglitazone as a candidate chemoprevention agent for lung cancer: A pilot window trial in early stage NSCLC. Journal of Clinical Oncology 32, no. 15_suppl (May 20, 2014) 1581-1581. DOI: 10.1200/jco.2014.32.15_suppl.1581 |
| 84 | Teramukai S, Kitano T, Kishida Y, Kawahara M, Kubota K, Komuta K, Minato K, Mio T, Fujita Y, Yonei T, Nakano K, Tsuboi M, Shibata K, Furuse K, Fukushima M. Pretreatment neutrophil count as an independent prognostic factor in advanced non-small-cell lung cancer: an analysis of Japan Multinational Trial Organisation LC00-03. Eur J Cancer. 2009 Jul;45(11):1950-8. doi: 10.1016/j.ejca.2009.01.023. |
| 85 | Seider MJ, Pugh SL, Langer C, Wyatt G, Demas W, Rashtian A, Clausen CL, Derdel JD, Cleary SF, Peters CA, Ramalingam A, Clarkson JE, Tomblyn M, Rabinovitch RA, Kachnic LA, Berk LB; NRG Oncology. Randomized phase III trial to evaluate radiopharmaceuticals and zoledronic acid in the palliation of osteoblastic metastases from lung, breast, and prostate cancer: report of the NRG Oncology RTOG 0517 trial. Ann Nucl Med. 2018 Oct;32(8):553-560. doi: 10.1007/s12149-018-1278-4. |
| 86 | Zhang M, Hong JA, Kunst TF, Bond CD, Kenney CM, Warga CL, Yeray J, Lee MJ, Yuno A, Lee S, Miettinen M, Ripley RT, Hoang CD, Gnjatic S, Trepel JB, Schrump DS. Randomized phase II trial of a first-in-human cancer cell lysate vaccine in patients with thoracic malignancies. Transl Lung Cancer Res. 2021 Jul;10(7):3079-3092. doi: 10.21037/tlcr-21-1. |
| 87 | T. Tsakiridis, G. Pond, J. Wright, P. Ellis, B.S. Abdulkarim, N. Ahmed, A.G. Robinson, M. Valdes, G. Okawara, A. Swaminath, M. Wierzbicki, M. Levine, Randomized Phase II Trial of Metformin in Combination with Chemoradiotherapy (CRT) in Locally Advanced Non-Small Cell Lung Cancer (LA-NSCLC) the OCOG-ALMERA trial (NCT02115464). International Journal of Radiation Oncology. (2020), 108 (Supple 3), S104 DOI:https://doi.org/10.1016/j.ijrobp.2020.07.2284 |
| 88 | Leung LS, Neal JW, Wakelee HA, Sequist LV, Marmor MF. Rapid Onset of Retinal Toxicity From High-Dose Hydroxychloroquine Given for Cancer Therapy. Am J Ophthalmol. 2015 Oct;160(4):799-805.e1. doi: 10.1016/j.ajo.2015.07.012. |
| 89 | Peters S, Danson S, Ejedepang D, Dafni U, Hasan B, Radcliffe HS, Bustin F, Crequit J, Coate L, Guillot M, Surmont V, Rauch D, Rudzki J, O'Mahony D, Barneto Aranda I, Scherz A, Tsourti Z, Roschitzki-Voser H, Pochesci A, Demonty G, Stahel RA, O'Brien M. Combined, patient-level, analysis of two randomised trials evaluating the addition of denosumab to standard first-line chemotherapy in advanced NSCLC - The ETOP/EORTC SPLENDOUR and AMGEN-249 trials. Lung Cancer. 2021 Nov;161:76-85. doi: 10.1016/j.lungcan.2021.09.002. |

**Supplementary Table S10:** List of articles identified and downloaded from clinicaltrials.gov

| 1 | Li KL, Li L, Zhang P, Kang J, Wang YB, Chen HY, He Y. A Multicenter Double-blind Phase II Study of Metformin With Gefitinib as First-line Therapy of Locally Advanced Non-Small-cell Lung Cancer. Clin Lung Cancer. 2017 May;18(3):340-343. doi: 10.1016/j.cllc.2016.12.003. |
| --- | --- |
| 2 | Nechushtan H, Hamamreh Y, Nidal S, Gotfried M, Baron A, Shalev YI, Nisman B, Peretz T, Peylan-Ramu N. A phase IIb trial assessing the addition of disulfiram to chemotherapy for the treatment of metastatic non-small cell lung cancer. Oncologist. 2015 Apr;20(4):366-7. doi: 10.1634/theoncologist.2014-0424. |
| 3 | Goss GD, Jonker DJ, Laurie SA, Weberpals JI, Oza AM, Spaans JN, la Porte C, Dimitroulakos J. A phase I study of high-dose rosuvastatin with standard dose erlotinib in patients with advanced solid malignancies. J Transl Med. 2016 Mar 31;14:83. doi: 10.1186/s12967-016-0836-6. |
| 4 | Reckamp KL, Krysan K, Morrow JD, Milne GL, Newman RA, Tucker C, Elashoff RM, Dubinett SM, Figlin RA. A phase I trial to determine the optimal biological dose of celecoxib when combined with erlotinib in advanced non-small cell lung cancer. Clin Cancer Res. 2006 Jun 1;12(11 Pt 1):3381-8. doi: 10.1158/1078-0432.CCR-06-0112. |
| 5 | Kelly K, Kittelson J, Franklin WA, Kennedy TC, Klein CE, Keith RL, Dempsey EC, Lewis M, Jackson MK, Hirsch FR, Bunn PA, Miller YE. A randomized phase II chemoprevention trial of 13-CIS retinoic acid with or without alpha tocopherol or observation in subjects at high risk for lung cancer. Cancer Prev Res (Phila). 2009 May;2(5):440-9. doi: 10.1158/1940-6207.CAPR-08-0136. |
| 6 | Han JY, Lee SH, Yoo NJ, Hyung LS, Moon YJ, Yun T, Kim HT, Lee JS. A randomized phase II study of gefitinib plus simvastatin versus gefitinib alone in previously treated patients with advanced non-small cell lung cancer. Clin Cancer Res. 2011 Mar 15;17(6):1553-60. doi: 10.1158/1078-0432.CCR-10-2525. |
| 7 | Marrone KA, Zhou X, Forde PM, Purtell M, Brahmer JR, Hann CL, Kelly RJ, Coleman B, Gabrielson E, Rosner GL, Ettinger DS. A Randomized Phase II Study of Metformin plus Paclitaxel/Carboplatin/Bevacizumab in Patients with Chemotherapy-Naïve Advanced or Metastatic Nonsquamous Non-Small Cell Lung Cancer. Oncologist. 2018 Jul;23(7):859-865. doi: 10.1634/theoncologist.2017-0465. |
| 8 | Skinner H, Hu C, Tsakiridis T, Santana-Davila R, Lu B, Erasmus JJ, Doemer AJ, Videtic GMM, Coster J, Yang AX, Lee RY, Werner-Wasik M, Schaner PE, McCormack SE, Esparaz BT, McGarry RC, Bazan J, Struve T, Paulus R, Bradley JD. Addition of Metformin to Concurrent Chemoradiation in Patients With Locally Advanced Non-Small Cell Lung Cancer: The NRG-LU001 Phase 2 Randomized Clinical Trial. JAMA Oncol. 2021 Sep 1;7(9):1324-1332. doi: 10.1001/jamaoncol.2021.2318. |
| 9 | Musallam KM, Taher AT. Re: Anti-angiogenic therapy using thalidomide combined with chemotherapy in small cell lung cancer: a randomized, double-blind, placebo-controlled trial. J Natl Cancer Inst. 2009 Dec 2;101(23):1657; author reply 1657-8. doi: 10.1093/jnci/djp371. |
| 10 | Gao HF, Liang Y, Zhou NN, Zhang DS, Wu HY. Aprepitant plus palonosetron and dexamethasone for prevention of chemotherapy-induced nausea and vomiting in patients receiving multiple-day cisplatin chemotherapy. Intern Med J. 2013 Jan;43(1):73-6. doi: 10.1111/j.1445-5994.2011.02637.x. |
| 11 | Hu Z, Cheng Y, Zhang H, Zhou C, Han B, Zhang Y, Huang C, Chang J, Song X, Liang J, Liang H, Bai C, Yu S, Chen J, Wang J, Pan H, Chitkara DK, Hille DA, Zhang L. Aprepitant triple therapy for the prevention of chemotherapy-induced nausea and vomiting following high-dose cisplatin in Chinese patients: a randomized, double-blind, placebo-controlled phase III trial. Support Care Cancer. 2014 Apr;22(4):979-87. doi: 10.1007/s00520-013-2043-9. |
| 12 | Garrido P, Pujol JL, Kim ES, Lee JM, Tsuboi M, Gómez-Rueda A, Benito A, Moreno N, Gorospe L, Dong T, Blin C, Rodrik-Outmezguine V, Passos VQ, Mok TS. Canakinumab with and without pembrolizumab in patients with resectable non-small-cell lung cancer: CANOPY-N study design. Future Oncol. 2021 Apr;17(12):1459-1472. doi: 10.2217/fon-2020-1098. |
| 13 | Zhou T, Zhang Y, Ma Y, Ma W, Wu X, Huang L, Feng W, Zhou H, Liu J, Zhao H, Zhang L, Yang Y, Huang Y. Comparison of aprepitant versus desloratadine for EGFR-TKI-induced pruritus: A randomized phase 2 clinical trial. Cancer. 2022 Nov 15;128(22):3969-3976. doi: 10.1002/cncr.34474. |
| 14 | Alite F, Balasubramanian N, Adams W, Surucu M, Mescioglu I, Harkenrider MM. Decreased Risk of Radiation Pneumonitis With Coincident Concurrent Use of Angiotensin-converting Enzyme Inhibitors in Patients Receiving Lung Stereotactic Body Radiation Therapy. Am J Clin Oncol. 2018 Jun;41(6):576-580. doi: 10.1097/COC.0000000000000324. |
| 15 | Koch A, Bergman B, Holmberg E, Sederholm C, Ek L, Kosieradzki J, Lamberg K, Thaning L, Ydreborg SO, Sörenson S; Swedish Lung Cancer Study Group. Effect of celecoxib on survival in patients with advanced non-small cell lung cancer: a double blind randomised clinical phase III trial (CYCLUS study) by the Swedish Lung Cancer Study Group. Eur J Cancer. 2011 Jul;47(10):1546-55. doi: 10.1016/j.ejca.2011.03.035. |
| 16 | Arrieta O, Barrón F, Padilla MS, Avilés-Salas A, Ramírez-Tirado LA, Arguelles Jiménez MJ, Vergara E, Zatarain-Barrón ZL, Hernández-Pedro N, Cardona AF, Cruz-Rico G, Barrios-Bernal P, Yamamoto Ramos M, Rosell R. Effect of Metformin Plus Tyrosine Kinase Inhibitors Compared With Tyrosine Kinase Inhibitors Alone in Patients With Epidermal Growth Factor Receptor-Mutated Lung Adenocarcinoma: A Phase 2 Randomized Clinical Trial. JAMA Oncol. 2019 Nov 1;5(11):e192553. doi: 10.1001/jamaoncol.2019.2553. |
| 17 | Harvey RD, Aransay NR, Isambert N, Lee JS, Arkenau T, Vansteenkiste J, Dickinson PA, Bui K, Weilert D, So K, Thomas K, Vishwanathan K. Effect of multiple-dose osimertinib on the pharmacokinetics of simvastatin and rosuvastatin. Br J Clin Pharmacol. 2018 Dec;84(12):2877-2888. doi: 10.1111/bcp.13753. |
| 18 | Chen Y, Li X, Zhang R, Xia Y, Shao Z, Mei Z. Effects of statin exposure and lung cancer survival: A meta-analysis of observational studies. Pharmacol Res. 2019 Mar;141:357-365. doi: 10.1016/j.phrs.2019.01.016. |
| 19 | Afzal MZ, Mercado RR, Shirai K. Efficacy of metformin in combination with immune checkpoint inhibitors (anti-PD-1/anti-CTLA-4) in metastatic malignant melanoma. J Immunother Cancer. 2018 Jul 2;6(1):64. doi: 10.1186/s40425-018-0375-1. |
| 20 | Gridelli C, Gallo C, Ceribelli A, Gebbia V, Gamucci T, Ciardiello F, Carozza F, Favaretto A, Daniele B, Galetta D, Barbera S, Rosetti F, Rossi A, Maione P, Cognetti F, Testa A, Di Maio M, Morabito A, Perrone F; GECO investigators. Factorial phase III randomised trial of rofecoxib and prolonged constant infusion of gemcitabine in advanced non-small-cell lung cancer: the GEmcitabine-COxib in NSCLC (GECO) study. Lancet Oncol. 2007 Jun;8(6):500-12. doi: 10.1016/S1470-2045(07)70146-8. |
| 21 | Bourigault P, Skwarski M, Macpherson RE, Higgins GS, McGowan DR. Investigation of atovaquone-induced spatial changes in tumour hypoxia assessed by hypoxia PET/CT in non-small cell lung cancer patients. EJNMMI Res. 2021 Dec 29;11(1):130. doi: 10.1186/s13550-021-00871-x. |
| 22 | Rosen LS, Gordon D, Tchekmedyian NS, Yanagihara R, Hirsh V, Krzakowski M, Pawlicki M, De Souza P, Zheng M, Urbanowitz G, Reitsma D, Seaman J. Long-term efficacy and safety of zoledronic acid in the treatment of skeletal metastases in patients with nonsmall cell lung carcinoma and other solid tumors: a randomized, Phase III, double-blind, placebo-controlled trial. Cancer. 2004 Jun 15;100(12):2613-21. doi: 10.1002/cncr.20308. |
| 23 | Chun SG, Liao Z, Jeter MD, Chang JY, Lin SH, Komaki RU, Guerrero TM, Mayo RC, Korah BM, Koshy SM, Heymach JV, Koong AC, Skinner HD. Metabolic Responses to Metformin in Inoperable Early-stage Non-Small Cell Lung Cancer Treated With Stereotactic Radiotherapy: Results of a Randomized Phase II Clinical Trial. Am J Clin Oncol. 2020 Apr;43(4):231-235. doi: 10.1097/COC.0000000000000632. |
| 24 | Tsakiridis T, Pond GR, Wright J, Ellis PM, Ahmed N, Abdulkarim B, Roa W, Robinson A, Swaminath A, Okawara G, Wierzbicki M, Valdes M, Levine M. Metformin in Combination With Chemoradiotherapy in Locally Advanced Non-Small Cell Lung Cancer: The OCOG-ALMERA Randomized Clinical Trial. JAMA Oncol. 2021 Sep 1;7(9):1333-1341. doi: 10.1001/jamaoncol.2021.2328. |
| 25 | Arrieta O, Varela-Santoyo E, Soto-Perez-de-Celis E, Sánchez-Reyes R, De la Torre-Vallejo M, Muñiz-Hernández S, Cardona AF. Metformin use and its effect on survival in diabetic patients with advanced non-small cell lung cancer. BMC Cancer. 2016 Aug 12;16:633. doi: 10.1186/s12885-016-2658-6. |
| 26 | Pandya KJ, Gajra A, Warsi GM, Argonza-Aviles E, Ericson SG, Wozniak AJ. Multicenter, randomized, phase 2 study of zoledronic acid in combination with docetaxel and carboplatin in patients with unresectable stage IIIB or stage IV non-small cell lung cancer. Lung Cancer. 2010 Mar;67(3):330-8. doi: 10.1016/j.lungcan.2009.04.020. |
| 27 | Arrieta O, Hernández-Pedro N, Maldonado F, Ramos-Ramírez M, Yamamoto-Ramos M, López-Macías D, Lozano F, Zatarain-Barrón ZL, Turcott JG, Barrios-Bernal P, Orozco-Morales M, Flores-Estrada D, Cardona AF, Rolfo C, Cacho-Díaz B. Nitroglycerin Plus Whole Intracranial Radiation Therapy for Brain Metastases in Patients With Non-Small Cell Lung Cancer: A Randomized, Open-Label, Phase 2 Clinical Trial. Int J Radiat Oncol Biol Phys. 2023 Mar 1;115(3):592-607. doi: 10.1016/j.ijrobp.2022.02.010. |
| 28 | Melosky B, Anderson H, Burkes RL, Chu Q, Hao D, Ho V, Ho C, Lam W, Lee CW, Leighl NB, Murray N, Sun S, Winston R, Laskin JJ. Pan Canadian Rash Trial: A Randomized Phase III Trial Evaluating the Impact of a Prophylactic Skin Treatment Regimen on Epidermal Growth Factor Receptor-Tyrosine Kinase Inhibitor-Induced Skin Toxicities in Patients With Metastatic Lung Cancer. J Clin Oncol. 2016 Mar 10;34(8):810-5. doi: 10.1200/JCO.2015.62.3918. |
| 29 | Owonikoko TK, Zhang G, Kim HS, Stinson RM, Bechara R, Zhang C, Chen Z, Saba NF, Pakkala S, Pillai R, Deng X, Sun SY, Rossi MR, Sica GL, Ramalingam SS, Khuri FR. Patient-derived xenografts faithfully replicated clinical outcome in a phase II co-clinical trial of arsenic trioxide in relapsed small cell lung cancer. J Transl Med. 2016 May 3;14(1):111. doi: 10.1186/s12967-016-0861-5. |
| 30 | Gore E, Bae K, Langer C, Extermann M, Movsas B, Okunieff P, Videtic G, Choy H. Phase I/II trial of a COX-2 inhibitor with limited field radiation for intermediate prognosis patients who have locally advanced non-small-cell lung cancer: radiation therapy oncology group 0213. Clin Lung Cancer. 2011 Mar;12(2):125-30. doi: 10.1016/j.cllc.2011.03.007. |
| 31 | Park H, Shapiro GI, Gao X, Mahipal A, Starr J, Furqan M, Singh P, Ahrorov A, Gandhi L, Ghosh A, Hickman D, Gallacher PD, Wennborg A, Attar EC, Awad MM, Das S, Dumbrava EE. Phase Ib study of eprenetapopt (APR-246) in combination with pembrolizumab in patients with advanced or metastatic solid tumors. ESMO Open. 2022 Oct;7(5):100573. doi: 10.1016/j.esmoop.2022.100573. |
| 32 | Gadgeel SM, Wozniak A, Ruckdeschel JC, Heilbrun LK, Venkatramanamoorthy R, Chaplen RA, Kraut MJ, Kalemkerian GP. Phase II study of docetaxel and celecoxib, a cyclooxygenase-2 inhibitor, in elderly or poor performance status (PS2) patients with advanced non-small cell lung cancer. J Thorac Oncol. 2008 Nov;3(11):1293-300. doi: 10.1097/JTO.0b013e31818b194e. |
| 33 | Rudin CM, Brahmer JR, Juergens RA, Hann CL, Ettinger DS, Sebree R, Smith R, Aftab BT, Huang P, Liu JO. Phase 2 study of pemetrexed and itraconazole as second-line therapy for metastatic nonsquamous non-small-cell lung cancer. J Thorac Oncol. 2013 May;8(5):619-23. doi: 10.1097/JTO.0b013e31828c3950. |
| 34 | Lee SM, James L, Buchler T, Snee M, Ellis P, Hackshaw A. Phase II trial of thalidomide with chemotherapy and as maintenance therapy for patients with poor prognosis small-cell lung cancer. Lung Cancer. 2008 Mar;59(3):364-8. doi: 10.1016/j.lungcan.2007.08.032. |
| 35 | Edelman MJ, Wang X, Hodgson L, Cheney RT, Baggstrom MQ, Thomas SP, Gajra A, Bertino E, Reckamp KL, Molina J, Schiller JH, Mitchell-Richards K, Friedman PN, Ritter J, Milne G, Hahn OM, Stinchcombe TE, Vokes EE; Alliance for Clinical Trials in Oncology. Phase III Randomized, Placebo-Controlled, Double-Blind Trial of Celecoxib in Addition to Standard Chemotherapy for Advanced Non-Small-Cell Lung Cancer With Cyclooxygenase-2 Overexpression: CALGB 30801 (Alliance). J Clin Oncol. 2017 Jul 1;35(19):2184-2192. doi: 10.1200/JCO.2016.71.3743. |
| 36 | Argiris A, Kut V, Luong L, Avram MJ. Phase I and pharmacokinetic study of docetaxel, irinotecan, and celecoxib in patients with advanced non-small cell lung cancer. Invest New Drugs. 2006 May;24(3):203-12. doi: 10.1007/s10637-005-3259-4. |
| 37 | Sanborn SL, Cooney MM, Dowlati A, Brell JM, Krishnamurthi S, Gibbons J, Bokar JA, Nock C, Ness A, Remick SC. Phase I trial of docetaxel and thalidomide: a regimen based on metronomic therapeutic principles. Invest New Drugs. 2008 Aug;26(4):355-62. doi: 10.1007/s10637-008-9137-0. |
| 38 | Schneider BJ, Kalemkerian GP, Kraut MJ, Wozniak AJ, Worden FP, Smith DW, Chen W, Gadgeel SM. Phase II study of celecoxib and docetaxel in non-small cell lung cancer (NSCLC) patients with progression after platinum-based therapy. J Thorac Oncol. 2008 Dec;3(12):1454-9. doi: 10.1097/JTO.0b013e31818de1d2. |
| 39 | Ai X, Song Z, Jian H, Zhou Z, Chen Z, Yu Y, Li Z, Lu S. Pyrotinib combined with thalidomide in advanced non-small-cell lung cancer patients harboring HER2 exon 20 insertions (PRIDE): protocol of an open-label, single-arm phase II trial. BMC Cancer. 2021 Sep 16;21(1):1033. doi: 10.1186/s12885-021-08759-8. |
| 40 | Reinmuth N, Meyer A, Hartwigsen D, Schaeper C, Huebner G, Skock-Lober R, Bier A, Gerecke U, Held CP, Reck M. Randomized, double-blind phase II study to compare nitroglycerin plus oral vinorelbine plus cisplatin with oral vinorelbine plus cisplatin alone in patients with stage IIIB/IV non-small cell lung cancer (NSCLC). Lung Cancer. 2014 Mar;83(3):363-8. doi: 10.1016/j.lungcan.2014.01.001 |
| 41 | Seider MJ, Pugh SL, Langer C, Wyatt G, Demas W, Rashtian A, Clausen CL, Derdel JD, Cleary SF, Peters CA, Ramalingam A, Clarkson JE, Tomblyn M, Rabinovitch RA, Kachnic LA, Berk LB; NRG Oncology. Randomized phase III trial to evaluate radiopharmaceuticals and zoledronic acid in the palliation of osteoblastic metastases from lung, breast, and prostate cancer: report of the NRG Oncology RTOG 0517 trial. Ann Nucl Med. 2018 Oct;32(8):553-560. doi: 10.1007/s12149-018-1278-4. |
| 42 | Albany C, Brames MJ, Fausel C, Johnson CS, Picus J, Einhorn LH. Randomized, double-blind, placebo-controlled, phase III cross-over study evaluating the oral neurokinin-1 antagonist aprepitant in combination with a 5HT3 receptor antagonist and dexamethasone in patients with germ cell tumors receiving 5-day cisplatin combination chemotherapy regimens: a hoosier oncology group study. J Clin Oncol. 2012 Nov 10;30(32):3998-4003. doi: 10.1200/JCO.2011.39.5558. |
| 43 | Li L, Jiang L, Wang Y, Zhao Y, Zhang XJ, Wu G, Zhou X, Sun J, Bai J, Ren B, Tian K, Xu Z, Xiao HL, Zhou Q, Han R, Chen H, Wang H, Yang Z, Gao C, Cai S, He Y. Combination of Metformin and Gefitinib as First-Line Therapy for Nondiabetic Advanced NSCLC Patients with EGFR Mutations: A Randomized, Double-Blind Phase II Trial. Clin Cancer Res. 2019 Dec 1;25(23):6967-6975. doi: 10.1158/1078-0432.CCR-19-0437. |
| 44 | Vishwanathan K, Dickinson PA, So K, Thomas K, Chen YM, De Castro Carpeño J, Dingemans AC, Kim HR, Kim JH, Krebs MG, Chih-Hsin Yang J, Bui K, Weilert D, Harvey RD. The effect of itraconazole and rifampicin on the pharmacokinetics of osimertinib. Br J Clin Pharmacol. 2018 Jun;84(6):1156-1169. doi: 10.1111/bcp.13534. |
| 45 | Mohamed AW, Elbassiouny M, Elkhodary DA, Shawki MA, Saad AS. The effect of itraconazole on the clinical outcomes of patients with advanced non-small cell lung cancer receiving platinum-based chemotherapy: a randomized controlled study. Med Oncol. 2021 Feb 9;38(3):23. doi: 10.1007/s12032-021-01475-0. |
| 46 | Scherpereel A, Berghmans T, Lafitte JJ, Colinet B, Richez M, Bonduelle Y, Meert AP, Dhalluin X, Leclercq N, Paesmans M, Willems L, Sculier JP; European Lung Cancer Working Party (ELCWP). Valproate-doxorubicin: promising therapy for progressing mesothelioma. A phase II study. Eur Respir J. 2011 Jan;37(1):129-35. doi: 10.1183/09031936.00037310. |
| 47 | Scagliotti GV, Kosmidis P, de Marinis F, Schreurs AJM, Albert I, Engel-Riedel W, Schallier D, Barbera S, Kuo HP, Sallo V, Perez JR, Manegold C. Zoledronic acid in patients with stage IIIA/B NSCLC: results of a randomized, phase III study. Ann Oncol. 2012 Aug;23(8):2082-2087. doi: 10.1093/annonc/mds128. |
