## Supplementary material for "A Systematic Review of Literatures: To Identify Non-Cancer Drugs Repurposed for Lung Cancer": Inclusion and Exclusion criteria

**Figure 1:** Inclusion and exclusion criteria

A) **PubMed search**

Exclusion criteria:

Aim is to obtain articles suggesting drug repurposing based on human clinical data.

1. Duplicates and non-English, inaccessible articles were excluded.
2. Articles on in-vitro and in-vivo models, in-silico studies, computational analysis, studies on COVID-19, pulmonary hypertension, asthma or fibrosis.
3. Any chemo drugs indicated or not for lung cancer were excluded.

Inclusion criteria:

Aim to obtain the non-chemo drug generic name.

1. Articles reporting or suggesting non-chemo drugs for lung cancer based on findings in human clinical trials were included.
2. Orphan drug or previously well-known non-chemo drug received approval for cancer post-registration of trial are included for further search.
B) **ClinicalTrials.gov search**

Aim is to obtain the trial result.

Exclusion criteria:

1. All pre-clinical, review articles or articles published on similar non-chemo drug efficacy in cancer patients posted in trial registry, and phase-I study results were excluded
2. Other reasons includes non-lung cancer patients recruited in the studies, non-randomized study design, relevant trial registration number not found, meta-analysis were excluded.

Inclusion criteria:

1. Articles containing the trial registration number.
2. Randomized placebo-controlled trials were finally included.
C) **Google Scholar search**

Aim is to obtain trial result

Exclusion criteria:

1. All review articles, phase-I and non-randomized studies were excluded.
2. Articles not containing trial registration number, pre-clinical studies, meta-analysis were excluded.

Inclusion criteria:

1. Articles containing trial registration number.
2. Randomized placebo-controlled trials were finally included.
