## Supplementary material for "A Systematic Review of Literatures: To Identify Non-Cancer Drugs Repurposed for Lung Cancer": Prisma 2020 flow chart

**Identification of studies via other methods**

**Identification of studies via databases and registers**

Records identified from: 83 drugs and ‘’Lung cancer’’ searched in,

Register: ClinicalTrials.gov (n = 284 trials (NCT#) for 61 non-chemo)

Records removed *before screening*:

Duplicate records removed (n = 2)

Records marked as ineligible by automation tools (n = 0)

Records removed for other reasons (n = 2 inaccessible and n=1 non-English)

Records identified from*:

PubMed (n = 513)

Articles on drugs not retrieved:

n = 23 drugs no trials available on cancer drugs in ClinicalTrials.gov

**Identification**

Trials / Articles not retrieved ClinicalTrials.gov:

Title not available in NCT# (n=117)

Irrelevant literatures (n=751)

Phase-I trials and other reasons = (n=35)

Reports sought for retrieval

articles found in NCT# from ClinicalTrials.gov (n = 798)

google scholar (n = 1199)

Records screened (n = 508)

Records excluded**

(Not Lung cancer n = 73)

Articles assessed for eligibility using NCT# in clinicaltrials.gov (n=12) and google scholar (n = 89 relevant studies found with NCT#)

Reports sought for retrieval

(n = 435)

Reports not retrieved

(COVID-19 studies = 48)

Trials excluded google scholar:

1) No published result of NCT# (n=167)
2) Irrelevant Articles excluded (n=1110)

**Screening**

Reports excluded:

Pre-clinical (n =196)

Computational studies (n = 91)

Others including articles on cancer drugs (n = 55)

Reports assessed for eligibility

(n = 387)

Articles excluded (google scholar):

1) Abstracts: (n =32)
2) Phase-I studies: (n=12 for 7 drugs)
3) Other reason + non-randomized: (n=16+7)

Published results (n = 12+22 = 34 studies, 6 duplicates) for (n=22) non-Chemo drugs. 2 NCT# contained study results included.

Non-chemo identified (n = 84 in 45 articles)

Studies included in review

(n = 5 + 28 articles)

Reports of included studies

(n = 5+30 reports =35)

**Included**

*Consider, if feasible to do so, reporting the number of records identified from each database or register searched (rather than the total number across all databases/registers).

**If automation tools were used, indicate how many records were excluded by a human and how many were excluded by automation tools.

*From:* Page MJ, McKenzie JE, Bossuyt PM, Boutron I, Hoffmann TC, Mulrow CD, et al. The PRISMA 2020 statement: an updated guideline for reporting systematic reviews. BMJ 2021;372:n71. doi: 10.1136/bmj.n71. For more information, visit: <http://www.prisma-statement.org/>
